# Kinematic signature of high-risk labored breathing revealed by novel signal analysis

**DOI:** 10.1101/2023.06.08.23291170

**Authors:** William B. Ashe, Brendan D. McNamara, Swet M. Patel, Julia N. Shanno, Sarah E. Innis, Camille J. Hochheimer, Andrew J. Barros, Ronald D. Williams, Sarah J. Ratcliffe, J. Randall Moorman, Shrirang M. Gadrey

## Abstract

Breathing patterns (respiratory kinematics) contain vital prognostic information. They report on a dimension of physiology that is not captured by conventional vital signs. But for an informative physiomarker to become clinically valuable, it must be measureable with ease, accuracy, and reproducibility. We sought to enable the quantitative characterization of respiratory kinematics at the bedside. Using inertial sensors, we analyzed upper rib, lower rib, and abdominal motion of 108 patients with respiratory symptoms during a hospital encounter (582 two-minute recordings). We measured the average respiratory rate and 33 other signal characteristics that had an explainable correspondence with clinically significant breathing patterns. K-means clustering revealed that the respiratory kinematic information was optimally represented by adding 3 novel measures to the average respiratory rate. We selected measures representing respiratory rate variability, respiratory alternans (rib-predominant breaths alternating with abdomen-predominant ones), and recruitment of accessory muscles (increased upper rib excursion). Latent profile analysis of these measures revealed a phenotype consistent with labored breathing. Poisson regression showed that the rate at which a patient’s recordings exhibited the labored breathing phenotype was significantly associated (p<0.01) with the severity of illness (discharge home v/s acute-care hospitalization v/s critical-care hospitalization). Notably, labored breathing was frequently detectable (21%) when the respiratory rate was normal, and it improved discrimination for critical illness. These findings validate the feasibility of respiratory kinematic phenotyping in routine healthcare settings, and demonstrate its clinical value. Further research into respiratory kinematic characteristics may reveal novel pathophysiologic mechanisms, advance the efficacy of predictive analytics, and enhance patient safety.

## INTRODUCTION

Breathing patterns (or respiratory kinematics) contain vital diagnostic and prognostic information. The normal breathing pattern is slow, regular cycles, usually 8-20 per minute, and a stable phenotype of predominantly abdominal and/or lower thoracic motion. These features make for an unmistakably effortless appearance. To clinicians, a normal breathing pattern is an important and reassuring physical examination finding *(1)*. In contrast, abnormal breathing patterns are important markers of disease *(2)*.

Labored breathing during acute illness, for example, is a major red-flag sign which often heralds clinical deterioration *(3)*; it can result from abnormalities of rate, rhythm, and relative movements of the upper chest, lower chest, and abdomen. Increased respiratory rate, or tachypnea, is an important feature of labored breathing. Its association with clinical deterioration is well known, as evidenced by its use in prominent criteria like quick Sequential Organ Failure Assessment (qSOFA) and National Early Warning Score (NEWS) *(4, 5)*. Additionally, the rhythm of labored breathing can be irregular and erratic *(6, 7)*. More subtle but equally important are abnormalities in the relative movements of the chest and abdomen. It is abnormal for the major motion of breathing to alternate between the chest and the abdomen; it signifies an overload of the primary respiratory muscles (diaphragm and intercostal) *(8)*. It is also abnormal for the upper chest to have large excursions during breaths; it signifies increased recruitment of accessory respiratory muscles in the neck (sternocleidomastoid and scalene) *(8)*.

Conventional vital signs do not report on respiratory kinematics beyond the average respiratory rate (3), and clinicians rely on qualitative visual inspections for a complete assessment of high risk breathing patterns *(9)*. Such assessments lack sensitivity and inter-rater reliability *(10)*, and they are manual effort intensive. Methods like plethysmography (inductance or optoelectronic) allow automated, quantitative analysis, but their hardware and/or calibration protocols are too cumbersome for everyday practice *(8, 11, 12)*.

We previously developed time-series methods to detect breath intervals and characterize respiratory kinematics using inertial sensors in healthy adults in an exercise physiology laboratory *(13)*. Here, we evaluated these respiratory kinematic characteristics at the hospital bedside. We describe a clinically explainable, high-dimensional signature of labored breathing, and validate it as a physiomarker of clinical deterioration during hospital encounters.

## MATERIALS AND METHODS

### Study design overview

At the University of Virginia medical center (UVAMC), we recruited 121 patients during their hospital encounters for shortness of breath. We recorded thoraco-abdominal kinematics using inertial sensors. We measured the respiratory rate and 33 other features of the inertial signals that had a clinically explainable correspondence with well-known breathing patterns. We used unsupervised k-means clustering to determine the number of measures that optimally represented our patients’ respiratory kinematics and selected one measure representing each cluster. To these representative measures, we applied latent profile analysis to identify respiratory kinematic phenotypes. Using Poisson regression, we studied the association between the respiratory kinematic phenotypes and severity of illness.

### Patient enrollment and signal recording

Between November 2020 and May 2021, we recruited 121 patients during their hospital encounter for shortness of breath. We used adulthood (age ≥ 18 years) and active respiratory symptoms as the inclusion criteria. We excluded patients with significant acute or chronic neuro-cognitive deficits that impaired their ability to follow commands. We excluded patients requiring non-invasive positive pressure ventilation (NIPPV) or invasive mechanical ventilation, since these can alter thoraco-abdominal respiratory kinematics. We recorded patients’ demographic (like age and sex), biometric (like body mass index and chest circumference), and clinical information (like chronic disease burden). We assessed the severity of their illness by recording the intensity of care they required in the span of 24 hours from the kinematic recording (discharge home vs non-critical care in hospital vs critical care in hospital). We defined critical care as admission to an intensive care unit (ICU) or requirement of ICU level interventions (intubation for mechanical ventilation, NIPPV, high FiO_2_ devices [FiO_2_: Fraction of inspired oxygen; high: >50%], or CPR [cardiopulmonary resuscitation]).

We used a network of 6 inertial sensors to record the kinematics of the upper rib-cage (midline sternal head and bilateral second rib in the midclavicular line), the lower rib-cage (bilateral eighth rib in the anterior axillary line), and the abdomen (midline abdomen near umbilicus). At each of the six locations, we analyzed three orthogonal axes each of accelerometer and gyroscope signals, making a total of 36 synchronized respiratory kinematic signals. We used a sampling frequency of 100Hz for all signals. Multiple two-minute recordings were obtained from each patient over a maximum of 6 hours. The number of recordings and the intervals between them varied depending on clinical circumstances and other factors (see section 1 of the supplementary file entitled “Prototype specifications”).

### Signal processing and motion artefact detection

We filtered all signals using a non-causal band-pass filter with corner frequencies of 0.05 Hz and 1 Hz. We used 4th and 6th order Butterworth kernels for the low and high filters, respectively; zero-phase filtering was achieved with a forward-backward filtering method, effectively doubling the strength of the filters to 8th and 12th orders.

To detect motion artefact, we combined the respiratory rate information in all 36 signal streams (6 sensors per patient; 6 inertial streams per sensor). First, we separated breaths within individual inertial signal streams using a method that we previously described *(13)*. The method uses the analytic representation, where a Hilbert transform of a signal is plotted on an imaginary *y*-axis against the untransformed signal on the real *x*-axis of a complex plane. The angle between a point on this representation and the real x-axis, with the origin as the vertex, is the phase angle of the wave *(13)*. For a quasi-cyclic process that is repetitive but not strictly periodic (like breathing), the phase angle tracks the same point on the signal across different cycles (e.g. peak or trough). Using these phase-based breath landmarks, we extracted respiratory rate time series just as heart rate time series are extracted from electrocardiograms using R-wave landmarks *(14)*. By repeating this process for each of the 36 streams of data per patient, we obtained 36 respiratory time series. In noise-free segments of the signal, a large proportion of the 36 rate series converged on the true respiratory rate. In noisy segments, there was no such convergence. This process is depicted in Figure 1 (for additional details, see section 2 of the supplementary file entitled “Detecting non-respiratory motion artefact”). We considered a segment of the signal as clean if the cluster of convergence included a majority (≥18) of the respiratory rates in that segment. We considered a 2-minute recording as acceptable for analysis if it contained at least 30 seconds of clean signal.

**Figure 1.**
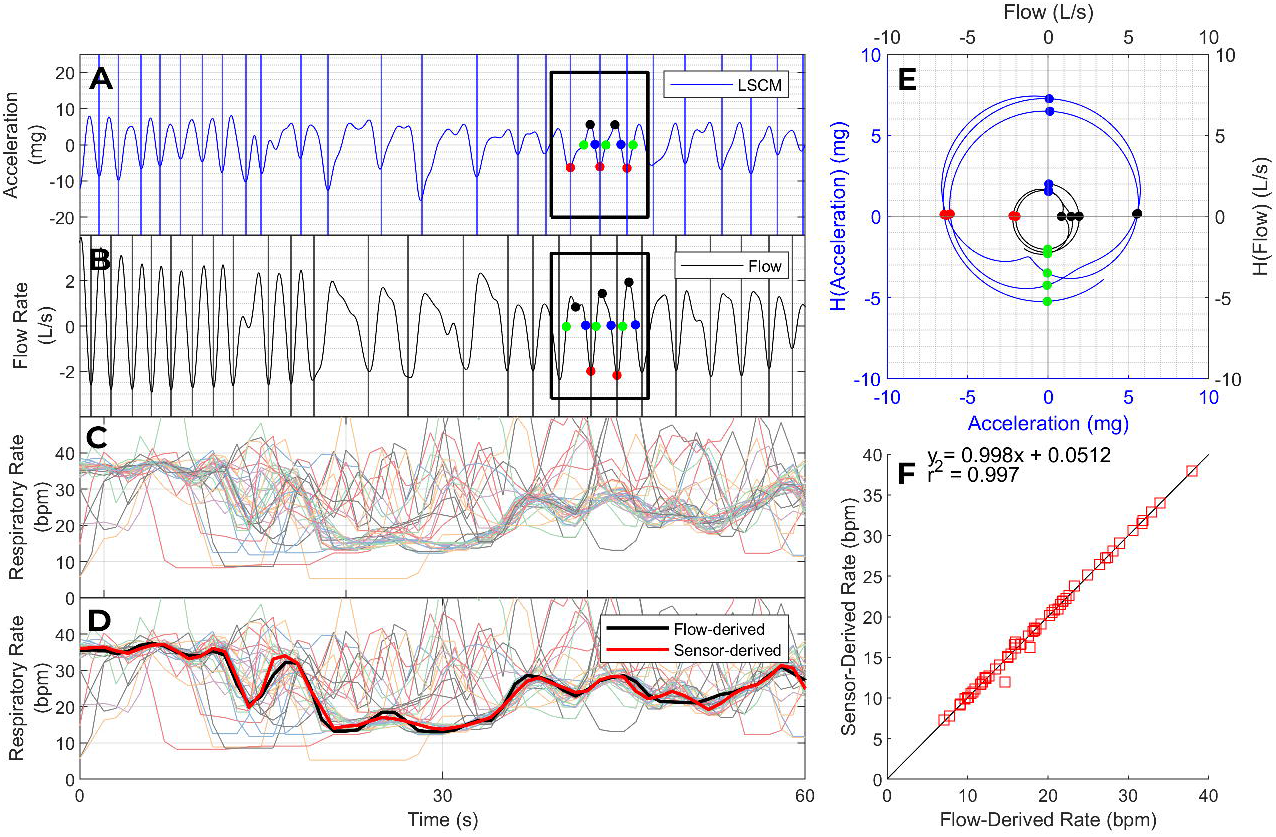
Respiratory rate and rhythm. Panels A and B show a respiratory kinematic signal from an accelerometer (A) and a synchronized volumetric air flow signal (B). The colored dots correspond to phase landmarks in the analytic representation of these signals (El, evenly spaced in phase at intervals of π/2 radians. The analytic representation of a signal is a complex plane where the Hilbert transform of a signal is represented on the imaginary y-axis as a function of the untransformed signal on the real x-axis. The phase of any point of the analytic signal is the angle between a line joining that point to the origin and the positive x-axis. We used phase landmarks to identify breath intervals and to extract an interpolated instantaneous respiratory rate series from each kinematic signal. Panel C shows 36 interpolated respiratory rate time series from one patient (6 kinematic signals per location; 6 locations). The rates extracted from clean kinematic signals converged on the true respiratory rate whereas the rates extracted from noisy kinematic signals deviated from the true respiratory rate to a varying degree. We identified the largest cluster of converging respiratory rates (thick bundle in Panel C) and adjudicated its centroid as our estimate of the respiratory rate (black line in Panel D). The final kinematics-derived rate series (black line) matched the flow-derived rate series (red line) with high fidelity. Panel F plots the kinematics-derived respiratory rate as a function of flow-derived respiratory rate in 60 paired recordings, with 95% limits of agreement of± 0.9 breaths per minute.

### Extraction of signal features and characterizing their time series dynamics

To evaluate the rate and rhythm of breathing we extracted a respiratory rate time series. As described above, we combined the information from all 36 respiratory rate time series by identifying a cluster of respiratory agreement. We used the centroid of this cluster as the final respiratory rate. We measured the central tendency and dispersion of the rate series. To externally validate this approach, we used respiratory kinematic signals that we had previously recorded in UVA’s exercise physiology lab (EPL) *(13)*. In this dataset, we had access to volumetric air flow signals (recorded using EPL equipment) that were synchronized with the kinematic signals. The signals were obtained in three exercise stages (rest, lactate threshold, and exhaustion) from 20 healthy subjects (i.e. 60 sets of synchronized flow and kinematic signals). We compared the kinematics-derived mean respiratory rates with the flow-derived mean respiratory rates using the Bland-Altman method (i.e. by calculating bias and 95% limit of agreement). And we calculated the cross-correlation coefficients between kinematics-derived and flow-derived rate time series. For details see section 3 of the supplementary file entitled “Validating respiratory rates in the exercise physiology laboratory”.

To measure the relative movements of the chest and abdomen, we extracted time series based on the instantaneous amplitude of linear acceleration signals. Once again, we relied on the analytic representation of the inertial signals. In this complex plane representation, the magnitude (distance from origin) of any point is the instantaneous amplitude of the corresponding point of the untransformed signal. We extracted features that represented (a) signal amplitude at individual sensor locations, (b) the average of the ratio of signal amplitude at one location to that in another location, and (c) the variability in the ratio of signal amplitude at one location to that in another location (For details see section 3 of the supplementary file entitled “Validating respiratory rates in the exercise physiology laboratory”).

In all, we extracted the respiratory rate and 33 measures based on a clinically explainable correspondence with well-known breathing motion patterns.

We selected mean respiratory rate for final statistical analysis since it is a kinematic metric with well-known clinical import. We applied k-means clustering to the remaining 33 metrics. We used the WSS plot (within sum of squares) to determine the number of clusters that best represented the observed respiratory kinematic variability. We used clinical domain expertise to select one metric from each cluster. This selection is detailed in section 5 of the supplementary file entitled “Optimal metric selection and latent profile analysis”.

### Statistical Analysis

We analyzed the scoring patterns across the selected respiratory kinematic measures using latent profile analysis (LPA) to uncover breathing pattern phenotypes. In this analysis, we used finite Gaussian mixture models estimated via the Expectation-Maximization (EM) algorithm *(15)*. We set initial values using hierarchical model-based agglomerative clustering, and we used Bayesian Information Criterion (BIC) to select optimal number of profile clusters (see section 5 of the supplementary file entitled “Optimal metric selection and latent profile analysis”). We identified the profile most consistent with the clinical construct of labored breathing. We used Poisson regression to model the association between the rate of the labored breathing phenotypes in each patient (number of recordings with labored breathing / total number of recordings) and the severity of illness (discharge home vs non-critical care in hospital vs critical care in hospital). We chose this approach to account for the variable number of measures per patient.

## RESULTS

### Signal quality and cohort characteristics

We successfully recorded 790 high-quality two-minute inertial signals from 108 of the 121 patients during their hospital encounter for shortness of breath. 582 (74%) of the 790 recordings met our acceptability criterion; at least 30 seconds of these 582 recordings was free of non-respiratory motion artefacts. Each of the 108 patients had at least 1 acceptable recording, and the median number of acceptable recordings per patient was 5 (range 1 - 11).

The median age of our patients was 65 years (range 19 - 91 years) and 54% were females. Consistent with the typical demographics of our medical center patients, 85% were White; 13% were Black, and 2% belonged to other racial groups. At least one chronic pulmonary condition was present in 42% of the patients (n = 46). Chronic obstructive pulmonary disease (n =35) and asthma (n=15) were the most common chronic conditions. Other diagnoses (n = 13) included interstitial lung disease, bronchiectasis, lung cancer, and sarcoidosis; some patients had more than one diagnosis. Twenty six patients required critical care hospitalization, 60 required acute-care hospitalization, and 22 were discharged home. Of the 582 recordings, 120 (21%) were from the 26 patients that required critical-care hospitalization, 355 (61%) were from the 60 patients that required acute-care hospitalizations, and 107 (18%) were from the 22 patients that were discharged home.

### Examples of signal analysis

Figure 1 illustrates our method of assessing the rate and rhythm of breathing. To optimize accuracy, we combined the respiratory rate information in 36 kinematic streams per recording (6 kinematic streams per sensor [three axes each of accelerometer and gyroscope signals]; 6 sensor locations per recording). We tested the accuracy of this method in the exercise physiology laboratory dataset (see section 3 of the supplementary file entitled “Validating respiratory rates in the exercise physiology laboratory”). Compared to the gold standard respiratory rate series (obtained using volumetric air flow sensors), the kinematics-derived respiratory rate series was highly accurate (bias: -0.02 breaths per minute; 95% limits of agreement: ±0.9 breaths per minute; average cross correlation coefficient: 0.89). To assess the rate of breathing, we measured the mean of the kinematics-derived respiratory rate series; to assess the rhythm of breathing, we measured the dispersion of the kinematics-derived respiratory rate series (standard deviation and coefficient of variation of the respiratory rate time series, and 9 other measures; see section 4 of the supplementary file entitled “Details of the 33 novel respiratory kinematic metrics”).

Figure 2 illustrates our method of assessing the relative movements of the upper chest, lower chest, and abdomen by analyzing the instantaneous amplitudes of linear acceleration. This method allowed us to assess: (a) the average magnitude of movement at a certain location of the torso (e.g. to what extent do the upper ribs move during breathing), (b) the average relationship between movements at two locations (e.g. on average, is breathing rib dominant or abdomen dominant or mixed), and the (c) the degree of variability in the relationship between movements at two locations (e.g. do rib dominant breaths alternate with abdomen dominant breaths).

**Figure 2.**
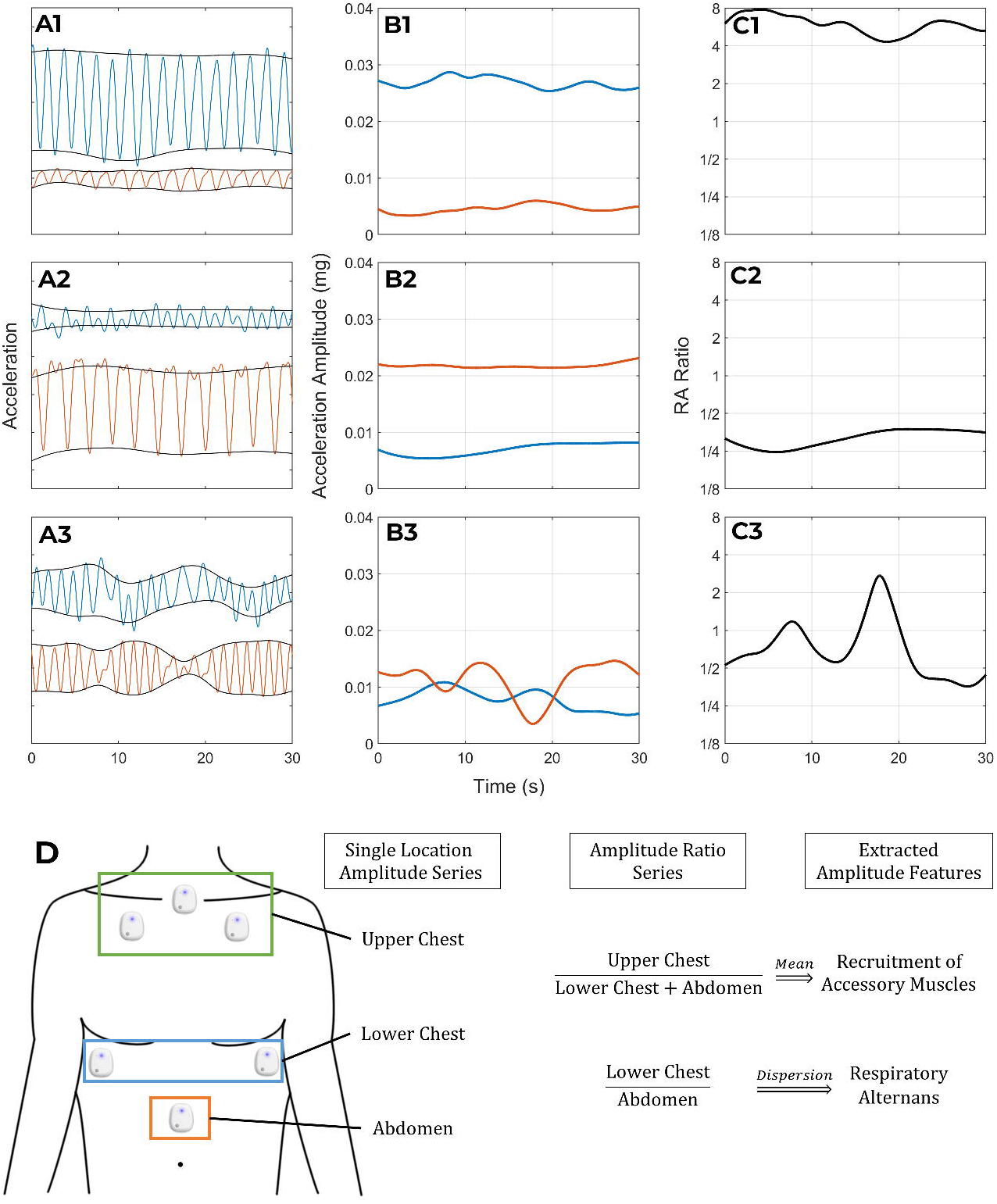
Relative movements of the chest and abdomen. This figure illustrates the information contained in the amplitude-relationships between the kinematic signals by displaying the signals of three patients (rows 1-3) whose breathing pattern varied in this regard. The first column of panels (A1-A3) shows one accelerometer signal from the lower rib (blue) and abdominal (orange) sensor of each patient. The instantaneous amplitude series of the signals shown in A1-A3 is plotted in panels B1-B3, respectively, and is derived from the magnitude (distance from origin) of the corresponding point in the analytic representation of that signal. Here, one dimension is plotted; calculations combined magnitudes from all three dimensions. Panels Cl-C3 show the amplitude ratio series that is obtained by dividing the lower rib amplitude series by the abdominal amplitude series in B1-B3. The first patient (Row 1) has a stable phenotype of rib-predominant breathing: the lower rib sensor (blue) has a higher amplitude kinematic signal than the abdominal sensor (orange) in Al; the lower rib (blue) has higher instantaneous amplitude than the abdomen (orange) in Bl; and the amplitude ratio series in Cl has a mean value greater than 1. In contrast to the rib-predominant breathing of the first patient, the second patient (Row 2) has abdomen-predominant breathing (A2 and B2) which results in an amplitude ratio that is less than 1 (C2). Despite their different breathing patterns, both of these patients have a stable pattern from one breath to the next. As a result, their amplitude ratio time series (Cl and C2) have low variability. In contrast, the third patient (row 3) has an unstable breathing pattern where rib predominant breaths alternate with abdomen predominant ones. This results in an amplitude ratio time series (C3) with high variability. Panel D schematically represents the derivation of our measures for respiratory alternans and recruitment of accessory muscles.

### A latent profile of labored breathing

Using signal analysis methods illustrated above, we extracted the average respiratory rate and 33 novel metrics from each recording: 11 metrics that reported on the rhythm of breathing, and 22 metrics that reported on the relative movements of the sensor locations (see section 4 of the supplementary file entitled “Details of the 33 novel respiratory kinematic metrics”). By applying k-means clustering *(16, 17)* to the 33 novel measures, we found that the respiratory kinematic variability across the 582 recordings was best described by three clusters of metrics (see section 5 of the supplementary file entitled “Optimal metric selection and latent profile analysis”). We selected one measure from each cluster using clinical domain expertise. These measures correspond with respiratory rate variability (short breaths alternating with long ones), respiratory alternans (rib-predominant breaths alternating with abdomen-predominant or mixed breaths), and recruitment of accessory muscles (increased upper chest expansion caused by contraction of neck muscles like scalene and sternocleidomastoid). These metrics were not correlated with each other; correlation coefficients were under 0.2 in all pairs.

We applied latent profile analysis*(18–20)* using the respiratory rate and the aforementioned three novel kinematic characteristics (respiratory rate variability, respiratory alternans, and recruitment of accessory muscles). Based on a plateau (“elbow”) in BIC improvement, we determined that the data were most consistent with two latent phenotypes of breathing (see section 5 of the supplementary file entitled “Optimal metric selection and latent profile analysis”).

One phenotype, had high means in all four metrics (respiratory rate: 24bpm [70th percentile]; respiratory rate variability: 6.8bpm [76th percentile]; respiratory alternans: 0.53 [74th percentile]; recruitment of accessory muscles: 1.9 [81st percentile]). This is consistent with the clinical construct of labored breathing. The other phenotype had lower mean values, consistent with unlabored breathing (respiratory rate: 20bpm [42nd percentile]; respiratory rate variability: 4.6 bpm [46th percentile]; respiratory alternans: 0.37 [45th percentile]; recruitment of accessory muscles: 1.1 [42nd percentile]).

### Clinical value of the labored breathing phenotype

Figures 3 to 5 contain six respiratory kinematic recordings along with a narration of their clinical context. These cases were selected to illustrate the various scenarios where quantitative characterization of respiratory kinematics may significantly enhance medical decision making. For ease of illustration, we displayed a single kinematic stream at each sensor location to convey the clinical constructs that are measured by the metrics.

**Figure 3.**
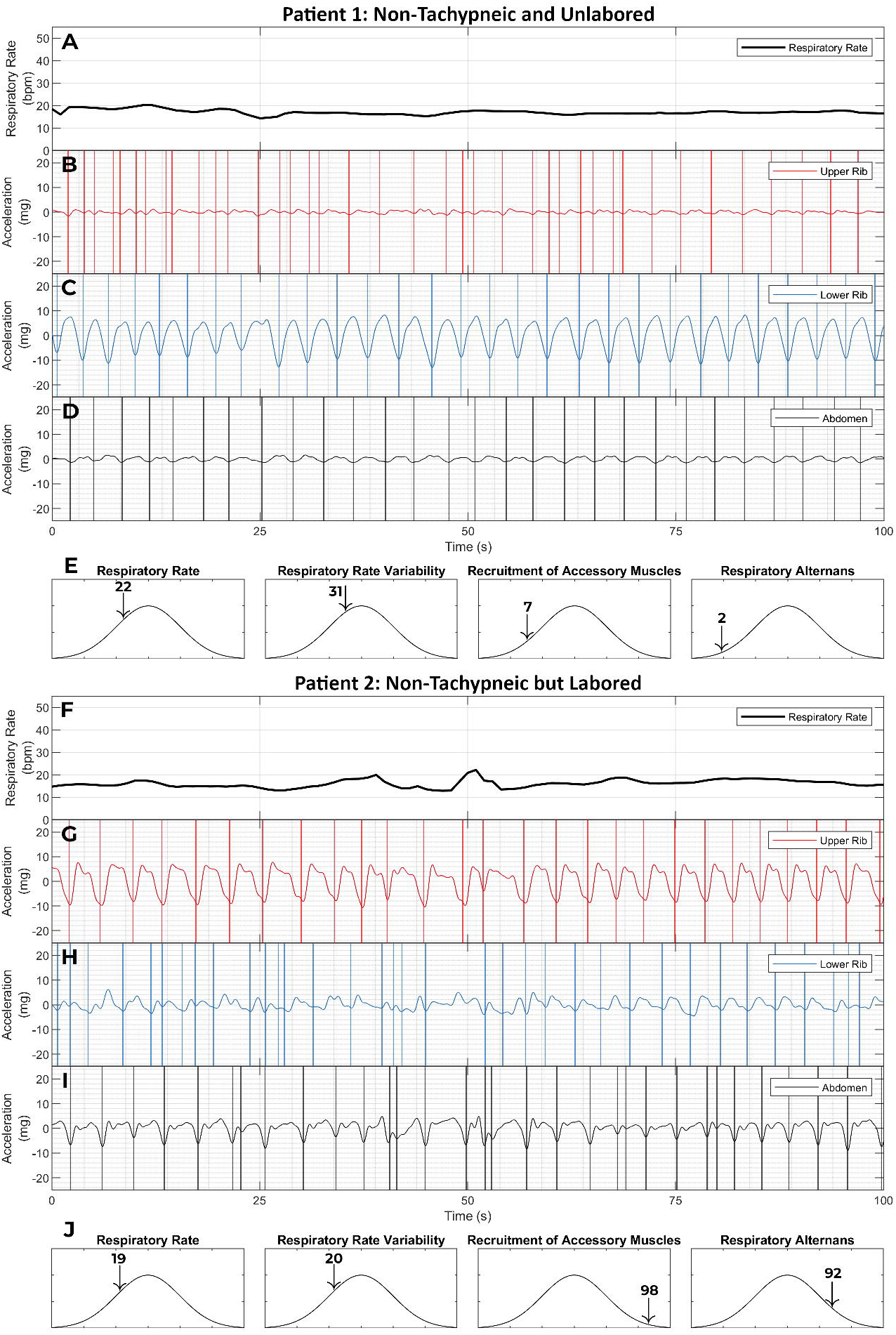
Labored breathing without tachypnea. This figure shows the respiratory kinematic recordings of two patients with a normal respiratory rate (≤20 breaths per minute). Patient 1 (panels A -E) was a 77 year old female who presented to the emergency room with shortness of breath. She exhibited not only a normal respiratory rate (22nd percentile in our dataset) but also an entirely unlabored breathing phenotype. Her breath intervals were regular. This was captured by a low respiratory rate variability (31st percentile). Her upper rib motion amplitude was low. This led to a low value on the recruitment of accessory muscles metric (7th percentile). And finally, she had a stable phenotype of lower rib dominant breathing. As a result, her respiratory alternans metric was also low (2nd percentile). Her emergency room workup was negative for any major acute illness. She was diagnosed with a viral upper respiratory infection and discharged home from the emergency room. Patient 2 (panels F - J) was a 57 year old male who presented with shortness of breath and lethargy. Patient 2 also had a normal respiratory rate (19th percentile) and a low respiratory rate variability (20th percentile). Yet, he was classified as having labored breathing. This was driven by the recruitment of accessory muscle and respiratory alternans metrics which were at the 99th and 92nd percentiles. The elevated respiratory alternans metric resulted from the variable abdominal signal amplitude relative to a constant lower rib amplitude. The elevated recruitment of accessory muscles metric corresponds with the fact that the amplitude of the upper rib signal exceeds that of the lower rib and/or abdominal signals. He was found to have acute on chronic hypoxemic and hypercarbic respiratory failure from severe chronic obstructive pulmonary disease. He required 15 liters per minute supplemental oxygen to maintain SpO2 > 88%; his pH was 7.2 and pCO2 was 105. He was initiated on corticosteroids, antibiotics, non-invasive positive pressure ventilation and admitted to the ICU (not intubated only due to his advanced directive). These examples show how a high-dimensional analysis of respiratory kinematics can improve discrimination for critical illness. [SpO2: oxygen saturation on pulse oximetry; pH: potential of hydrogen; pCO2: partial pressure of carbon dioxide; ICU: intensive care unit]

Figure 3 contains the respiratory kinematic recordings of two patients who presented with shortness of breath. Neither patient was tachypneic; both had a normal and virtually identical respiratory rate. However, only one of these patients exhibited a labored breathing phenotype. The patient with non-tachypneic and non-labored breathing was uneventfully discharged home. The patient with the non-tachypneic but labored breathing was admitted to the intensive care unit for life-threatening respiratory failure. This vignette demonstrates how high-risk labored breathing is detectable even in the absence of tachypnea.

Figure 4 also contains the respiratory kinematic recordings of two patients who were initially admitted to the acute care for the treatment of pneumonia and mild hypoxemic respiratory failure. Both patients were tachypneic, and the risk predicted by the Pneumonia Severity Index was comparable *(21)*. However, the breathing patterns of one patient were more labored than the other (larger elevations in respiratory rate variability, recruitment of accessory muscles, and respiratory alternans). The patient with lower degrees of labored breathing derangements remained stable and was uneventfully discharged home after recovery. The patient with higher degrees of labored breathing derangements developed an abrupt clinical deterioration requiring an emergent endotracheal intubation and an unplanned transfer to the intensive care unit for life-threatening respiratory failure. That patient died within 24 hours of their respiratory kinematic recording. This vignette exemplifies the clinical scenario where respiratory kinematic characteristics may provide alerts that are more specific than tachypnea for respiratory muscle overload and imminent respiratory deterioration.

**Figure 4.**
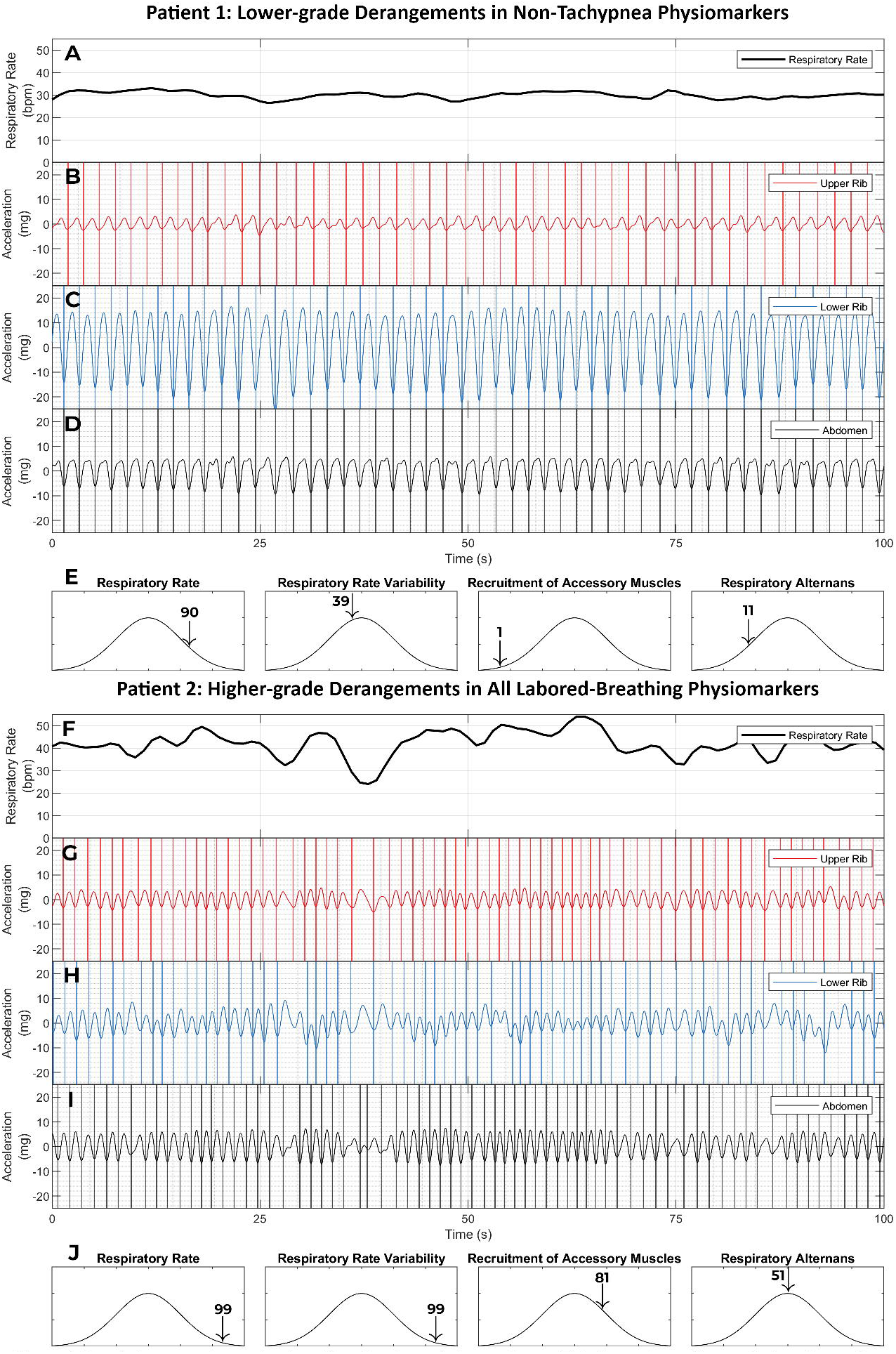
Respiratory muscle overload and imminent respiratory collapse. This figure shows the respiratory kinematic recordings of two female patients with elevated respiratory rates (90th and 99th percentiles for our dataset). They were both initially admitted to the acute care unit of the hospital for pneumonia and sepsis with acute respiratory failure. At admission, their supplemental oxygen needs were low (4 and 2 liters per minute) and their blood gas studies were normal (pH of 7.43 and 7.44; pCO2 of 33 and 40 mmHg). Their Pneumonia Severity Index scores were also comparable at 127 and 125 [scores of 91 -130 are Risk Class 4 (moderate risk) and are associated with a 30-day mortality of 9%]. Apart from tachypnea and hypoxemia, the drivers of risk were age, hyponatremia, and confusion in Patient 1, and fevers, anemia, and neoplastic disease (acute myelogenous leukemia) in patient 2. Despite the similarities in these patients’ presentations and conventional risk levels, their clinically trajectories were dramatically different. Patient 1 remained stable on the acute care unit after her kinematic recording and was discharged home after a 7 day hospitalization. In contrast, Patient 2 developed a sudden respiratory collapse within 6 hours of her kinematic recording (required emergent intubation), and died within 24 hours of this recording. Severe hypoventilation from respiratory muscle overload was deemed to be the likely mechanism of the collapse; this conclusion was consistent with the rapidity of collapse, unresponsiveness to oxygen supplementation prior to intubation (low Sp02 despite 100% FiO2), and acute hypercapnia (pH 6.9 mm Hg; pCO2 103mm Hg). A majority of both patients’ recordings were classified as labored breathing in the latent profile analysis (3 of 4 recordings in patient 1, and 3 of 3 recordings in patient 2). However, a comparison of the respiratory kinematic characteristics of these patients revealed an important difference. Patient 1 (the survivor) had lower grade derangements in all non-tachypnea physiomarkers than the deceased patient 2. The breath intervals were more regular in Panels B-D than in panels G-1 (respiratory rate variability at 39th percentile in panel E vs 99th percentile in panel J). The upper rib motion amplitude was much smaller than lower rib and/or abdominal motion amplitude in panels B-0 than in panels G-1(recruitment of accessory muscles at 1st percentile in panel G vs 81st percentile in panel J). And the breath phenotype was much more stable. Each breath had a same degree of lower rib dominance in panels C-0, whereas rib dominant breaths alternated with abdomen dominant ones in panels H-1(respiratory alternans at 11th percentile in panel E vs 51st percentile in panel J). Consistent with the conventional wisdom in the field of bedside physical diagnosis, these findings suggest that respiratory kinematic characteristics contain signatures that are much more specific for respiratory muscle overload than just tachypnea.

Figure 5 contains two respiratory kinematic recordings obtained from one asthmatic patient whose acute respiratory failure resolved rapidly after breathing treatments were administered. The recordings show improvements in the patient’s respiratory kinematic characteristics that match their clinical trajectory. This vignette highlights how respiratory kinematic characteristics can reveal dynamic pathophysiologic trends *(22)*.

**Figure 5.**
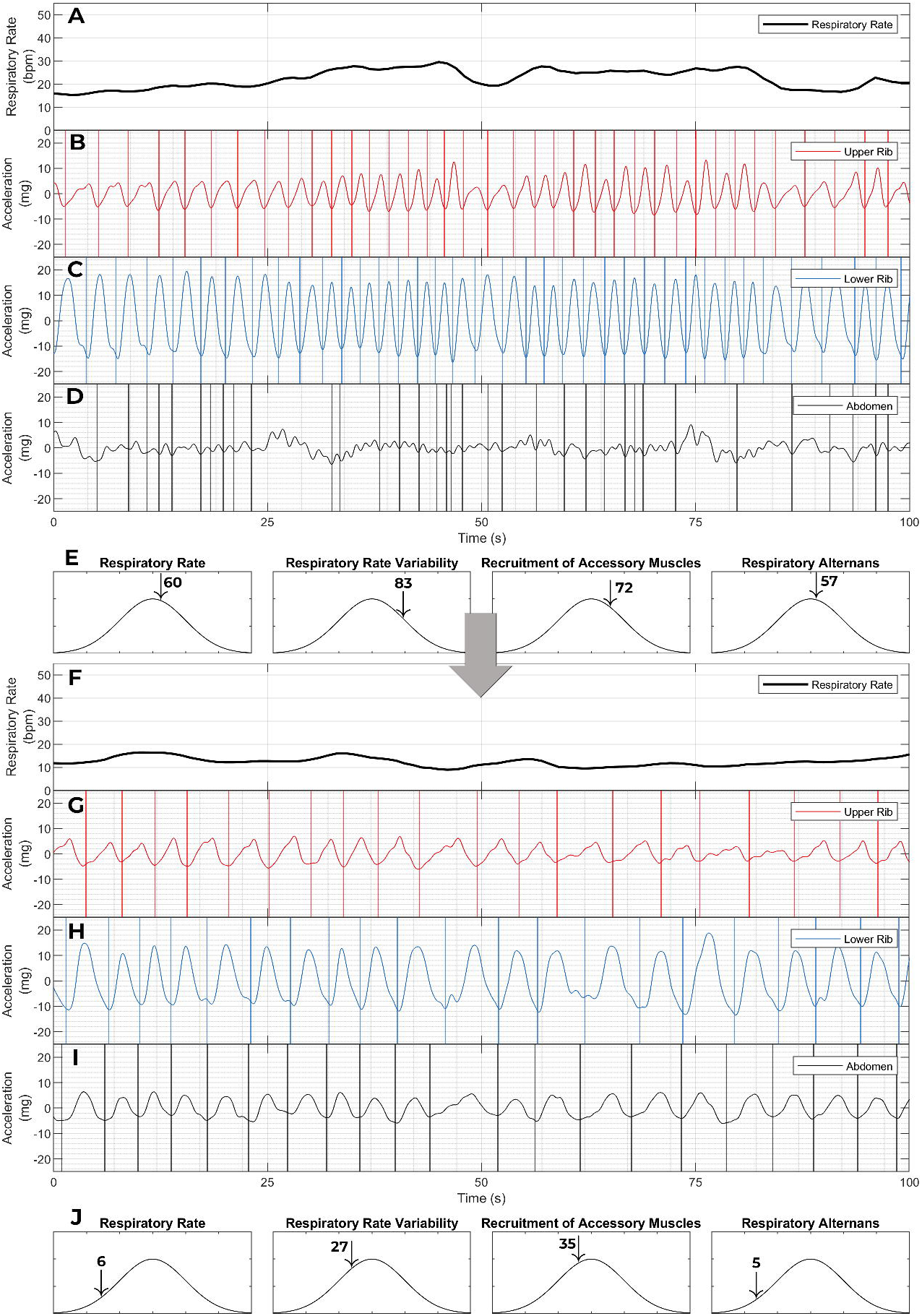
Personalized respiratory kinematic profiles and their trajectories. This figure contains two respiratory kinematic recordings of a 20 year old male who presented with common cold symptoms and shortness of breath. He was diagnosed with acute hypoxemic respiratory failure (required up to 10 liters per minute of supplemental oxygen) from an acute exacerbation of his mild intermittent asthma precipitated by a rhinovirus infection. With steroids and bronchodilator treatments, his hypoxemic respiratory failure rapidly resolved; his oxygen supplementation was stopped within 12 hours of treatment and he was discharged after a 48 hour acute care hospitalization. Only one (the first) of his four recordings was classified as labored breathing in our unsupervised latent profile analysis. Trends consistent with the clinical trajectory can be seen in the two recordings shown here. In the initial recording (top) patient’s breath intervals are shorter and more irregular than in the in the later recording (bottom). This corresponds with an improvement in our measures for respiratory rate (22 breaths per minute [60th percentile] to 12 breaths per minute [6th percentile]), and respiratory rate variability [83rd percentile to 27th percentile]. Note also, how the amplitude of upper rib signal improves (recruitment of accessory muscles metric goes from 72nd percentile to 35th percentile). Interestingly, this corresponds with an increase in the amplitude of the abdominal signal (In Panel D, there is virtually no organized abdominal signal; In Panel I, the abdominal signal is organized and its amplitude is higher than the upper rib). During an asthma exacerbation, dynamic hyperinflation can flatten the diaphragm and impair its contraction (20). Reduction in hyperinflation in response to treatment is a plausible explanation for such a change in kinematic recordings. This case highlights the potential for personalized respiratory kinematic profiling, whereby dynamic trends that deviate from an individual’s own baseline profile may provide early warnings of acute illness.

The overall frequency of the labored breathing phenotype was 33% (193 of 582 total recordings). Poisson regression revealed an association between the proportion of a patient’s recordings that contained the labored breathing phenotype, and the severity of the patient’s illness. The frequency of the labored breathing phenotype was lowest (21%) in patients who were discharged home. Compared to this group, the frequency of labored breathing was higher in patients who required non-critical care hospitalization (33% v/s 21%; p = 0.06) and in patients requiring critical care hospitalization (44% v/s 21%; p = 0.004). Further, as shown in Figure 6, the discrimination for critical illness improved when the labored breathing phenotype was added to tachypnea as a marker of respiratory distress.

**Figure 6.**
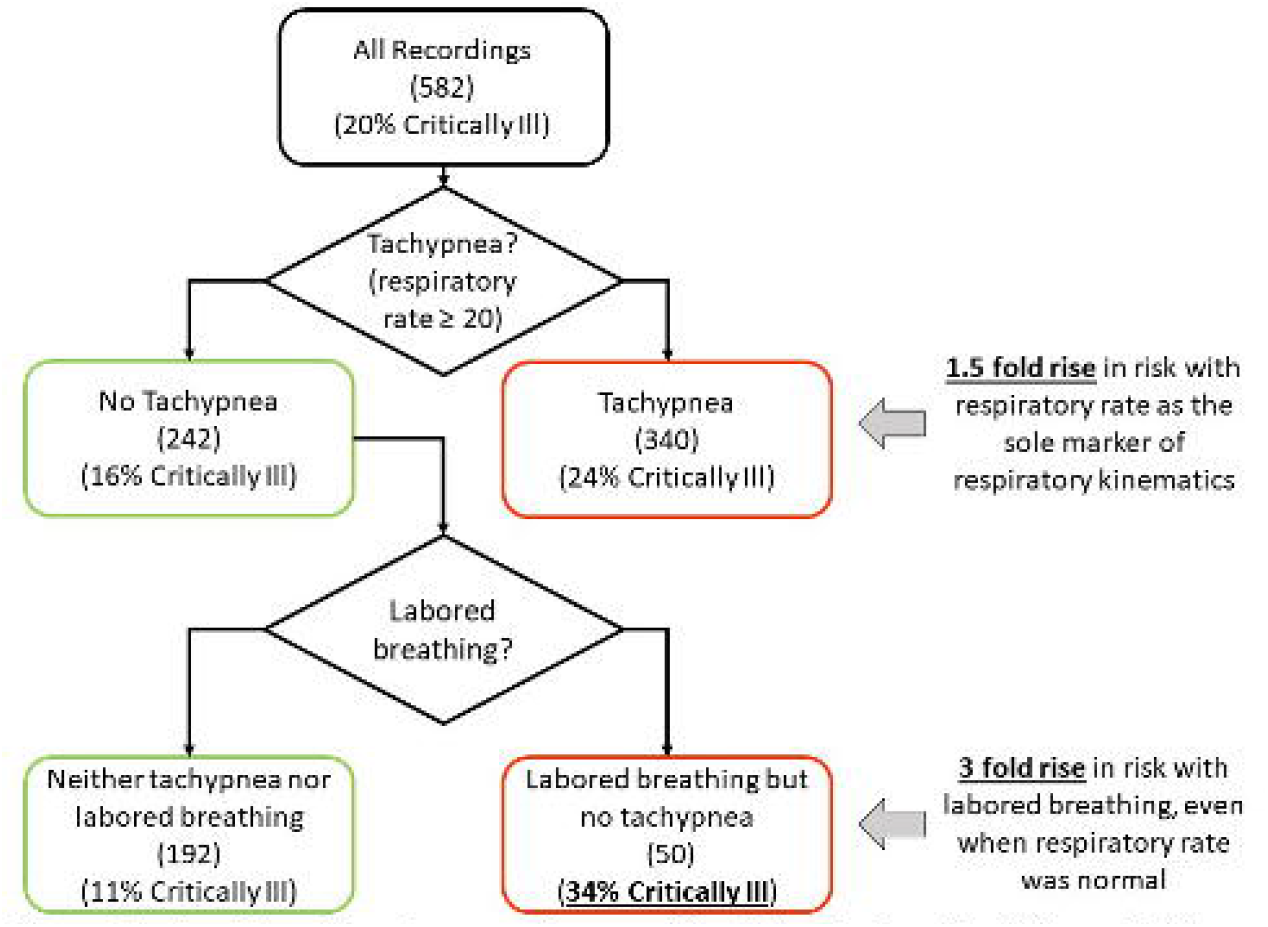
Labored breathing phenotype improved discrimination for critical illness. Multi-dimensional analysis of respiratory kinematics revealed a signature of labored breathing. On Poisson regression, the association of this signature with the severity of illness was statistically significant. The clinical significance of this finding is conveyed by this figure. Of the 582 recordings, 20% belonged to patients who required critical care hospitalization. When we used respiratory rate as the sole respiratory kinematic physiomarker (which is the prevailing clinical standard), tachypnea (respiratory rate ≥ 20 breaths per minute) was associated with a 1.5 fold rise in risk of critical illness (24% vs. 16%). Arguably, this is a clinically useful degree of discrimination and affirms the validity of the kinematics-derived respiratory rate. Importantly, however, the discrimination improved when the low-risk (non-tachypneic) recordings were further classified based on the presence of the labored breathing. More than one fifth of the 242 non-tachypneic recordings contained the labored breathing phenotype. These recordings were associated with a 3 fold rise in risk of critical illness when compared to the non-tachypneic non-labored recordings (34% vs. 11%).

## DISCUSSION

We studied the breathing motion patterns of patients with active respiratory symptoms. Our major findings are that (a) respiratory kinematics are a rich source of physiological information, with more complexity than is adequately represented by the respiratory rate alone, and (b) high-risk respiratory kinematic phenotypes can be identified in an everyday clinical context through multi-dimensional analysis of respiratory kinematics.

Clinicians have long recognized that the respiratory rate is an important vital sign. Equally well understood is the degree of inaccuracy with which it is currently measured *(23, 24)*. We derived a method for respiratory rate estimation in our hospital patient population and validated that it results in highly accurate estimates in new populations (bias and 95% limit of agreement of -0.02 ± 0.9 breaths per minute in the exercise physiology laboratory subjects). Improving the accuracy of respiratory rate documentation can enhance patient safety, especially in the more remote parts of the world. For example, approximately 750,000 children under 5 years of age die from pneumonia every year *(25)*. UNICEF estimates that many deaths can be prevented if parents and community health workers can simply be trained to measure the respiratory rate and detect tachypnea *(26)*. But even this basic assessment (fast vs normal respiratory rate) cannot be reliably achieved in many parts of the world *(27)*. The respiratory rate estimation methods described in this paper may improve diagnosis and save lives.

Notwithstanding the importance of the respiratory rate, it is only one among many breathing motion patterns that convey respiratory distress. A patient’s breathing can exhibit high-risk labored breathing despite a normal respiratory rate. In such scenarios, the risk can be captured by detecting other abnormal patterns like excessive upper rib excursion during breathing or an unstable breath phenotype (chest predominant breaths alternating with abdomen predominant ones). Seasoned clinicians caution their trainees, therefore, against the use of respiratory rate as the sole marker of labored breathing *(28)*. In the absence of quantitative clinical and/or research tools, however, such insights have been largely restricted to the domain of conventional clinical wisdom. A major contribution of our study is to confirm this clinical intuition and provide one of the first estimates of its magnitude in a real-world clinical setting. Our novel physiomarkers revealed signs of labored breathing in a fifth of the recordings that had a normal respiratory rate, even when we used one of the most sensitive clinical definitions of tachypnea (respiratory rate ≥20 breaths per minute). The resulting reclassification led to improved discrimination for critical illness. This points to a major opportunity for improvement in clinical risk stratification and early warning scores. Unanticipated respiratory compromise and unplanned intubation are common and catastrophic events in patients hospitalized in general medical and surgical units of a hospital *(29, 30)*. Early warnings triggered by high-risk respiratory kinematic phenotypes may empower clinicians to address this menacing problem.

A major strength of this work is that it builds on principles that are fundamental to bedside diagnosis and to the pathophysiology of human ventilation. The physiomarker extraction was clinically-driven. As such, the respiratory kinematic characteristics reported in this study have an easily explainable correspondence with physical examination signs that are commonly used by clinicians. Additional clinically-driven kinematic signal analysis can lead to a more comprehensive characterization of respiratory kinematics. Thoraco-abdominal asynchrony (“abdominal paradox”), Cheyne-Stokes breathing, and apneas are examples of well-known physical examination findings that were not operationalized in this study. Moreover, a data-driven approach to physiomarker extraction may also be useful when larger respiratory kinematic datasets become available. In this latter approach, physiomarkers selection would be based on either an association with clinical outcomes (supervised learning), or an observed pattern in unlabeled data (unsupervised learning). Such physiomarkers may be poorly understood at first, but they may generate hypotheses and lead to the discovery of new pathophysiologic mechanisms. Another strength of our work is that acceptable recordings were unobtrusively achieved in a busy hospital environment. Every patient had at least 1 acceptable recording and 75% of all recordings were acceptable. This demonstrates the technical feasibility of large scale respiratory kinematic monitoring in real-world clinical settings.

This study is limited in generalizability because of convenience sampling. Also, due to the small sample size the study was not adequately powered to accommodate multivariable regression to adjust for variables other than respiratory kinematics (such as oxygenation, hemodynamics, and organ dysfunction). Here, we demonstrate the technical feasibility and clinical utility of a novel approach to respiratory physiological monitoring.

## Data Availability

All data produced in the present study are available upon reasonable request to the authors

## Acknowledgments

We thank Dr. Mark Sochor, Dr. Thomas Hartka, Ashley Simpson, Anjali Kapil, and Alexander Schwartz of UVA’s Emergency Medicine Research Office for their invaluable help with patient enrollment and data collection.

We thank Sharon Krueger, program director of the Ivy Foundation’s Biomedical Innovation Fund for her advice and support.

We thank David Chen, director of UVA’s Coulter Translational Research Partnership, for his role in establishing and supporting this multidisciplinary collaboration.

## Author contributions

Conceptualization: SMG, WBA, RDW, SJR, JRM

Methodology: SMG, WBA, AJB, RDW, SJR, JRM

Investigation: SMG, WBA, BDM, SMP, JNS, SEI, CJH

Visualization: SMG, WBA

Funding acquisition: SMG, WBA, RDW, SJR, JRM

Project administration: SMG

Supervision: SMG, AJB, RDW, SJR, JRM

Writing – original draft: SMG, WBA

Writing – review & editing: SMG, WBA, AJB, RDW, SJR, JRM

## References and Notes

1. M. J. Tobin, T. S. Chadha, G. Jenouri, S. J. Birch, H. B. Gazeroglu, M. A. Sackner, Breathing patterns. 1. Normal subjects. Chest 84, 202–205 (1983).

2. M. J. Tobin, T. S. Chadha, G. Jenouri, S. J. Birch, H. B. Gazeroglu, M. A. Sackner, Breathing patterns. 2. Diseased subjects. Chest 84, 286–294 (1983).

3. A. Tulaimat, R. M. Gueret, M. F. Wisniewski, J. Samuel, Association Between Rating of Respiratory Distress and Vital Signs, Severity of Illness, Intubation, and Mortality in Acutely Ill Subjects. Respiratory Care 59, 1338–1344 (2014).

4. M. Singer, C. S. Deutschman, C. W. Seymour, M. Shankar-Hari, D. Annane, M. Bauer, R. Bellomo, G. R. Bernard, J.-D. Chiche, C. M. Coopersmith, R. S. Hotchkiss, M. M. Levy, J. C. Marshall, G. S. Martin, S. M. Opal, G. D. Rubenfeld, T. van der Poll, J.-L. Vincent, D. C. Angus, The Third International Consensus Definitions for Sepsis and Septic Shock (Sepsis-3). JAMA 315, 801–810 (2016).

5. G. B. Smith, D. R. Prytherch, P. Meredith, P. E. Schmidt, P. I. Featherstone, The ability of the National Early Warning Score (NEWS) to discriminate patients at risk of early cardiac arrest, unanticipated intensive care unit admission, and death. Resuscitation 84, 465–470 (2013).

6. D. Garrido, J. J. Assioun, A. Keshishyan, M. A. Sanchez-Gonzalez, B. Goubran, Respiratory Rate Variability as a Prognostic Factor in Hospitalized Patients Transferred to the Intensive Care Unit. Cureus 10, e2100.

7. A. J. E. Seely, A. Bravi, C. Herry, G. Green, A. Longtin, T. Ramsay, D. Fergusson, L. McIntyre, D. Kubelik, D. E. Maziak, N. Ferguson, S. M. Brown, S. Mehta, C. Martin, G. Rubenfeld, F. J. Jacono, G. Clifford, A. Fazekas, J. Marshall, Canadian Critical Care Trials Group (CCCTG), Do heart and respiratory rate variability improve prediction of extubation outcomes in critically ill patients? Crit Care 18, R65 (2014).

8. American Thoracic Society/European Respiratory Society, ATS/ERS Statement on respiratory muscle testing. Am. J. Respir. Crit. Care Med. 166, 518–624 (2002).

9. A. Tulaimat, W. E. Trick, DiapHRaGM: A mnemonic to describe the work of breathing in patients with respiratory failure. PLoS One 12 (2017), doi:10.1371/journal.pone.0179641.

10. A. Tulaimat, A. Patel, M. Wisniewski, R. Gueret, The validity and reliability of the clinical assessment of increased work of breathing in acutely ill patients. Journal of Critical Care 34, 111–115 (2016).

11. P. Laveneziana, A. Albuquerque, A. Aliverti, T. Babb, E. Barreiro, M. Dres, B.-P. Dubé, B. Fauroux, J. Gea, J. A. Guenette, A. L. Hudson, H.-J. Kabitz, F. Laghi, D. Langer, Y.-M. Luo, J. A. Neder, D. O’Donnell, M. I. Polkey, R. A. Rabinovich, A. Rossi, F. Series, T. Similowski, C. M. Spengler, I. Vogiatzis, S. Verges, ERS statement on respiratory muscle testing at rest and during exercise. Eur Respir J 53, 1801214 (2019).

12. M. J. Tobin, Breathing pattern analysis. Intensive Care Med 18, 193–201 (1992).

13. W. B. Ashe, S. E. Innis, J. N. Shanno, C. J. Hochheimer, R. D. Williams, S. J. Ratcliffe, J. R. Moorman, S. M. Gadrey, Analysis of respiratory kinematics: a method to characterize breaths from motion signals. Physiol. Meas. 43, 015007 (2022).

14. R. D. Berger, S. Akselrod, D. Gordon, R. J. Cohen, An efficient algorithm for spectral analysis of heart rate variability. IEEE Trans Biomed Eng 33, 900–904 (1986).

15. A. P. Dempster, N. M. Laird, D. B. Rubin, Maximum Likelihood from Incomplete Data via the EM Algorithm. Journal of the Royal Statistical Society. Series B (Methodological) 39, 1–38 (1977).

16. E. Forgy, Cluster analysis of multivariate data: efficiency versus interpretability of classifications. Biometrics. 21 (3): 768–769. JSTO^_J^R 2528559 (1965).

17. J. MacQueen, Some methods for classification and analysis of multivariate observations. Proceedings of 5th Berkeley Symposium on Mathematical Statistics and Probability. Vol. 1. University of California Press. 5.1, 281–298 (1967).

18. W. A. Gibson, Three multivariate models: Factor analysis, latent structure analysis, and latent profile analysis. Psychometrika 24, 229–252 (1959).

19. D. Oberski, in Modern Statistical Methods for HCI, Human–Computer Interaction Series. J. Robertson, M. Kaptein, Eds. (Springer International Publishing, Cham, 2016), pp. 275–287.

20. L. Scrucca, M. Fop, T. B. Murphy, A. E. Raftery, mclust 5: Clustering, Classification and Density Estimation Using Gaussian Finite Mixture Models. R J 8, 289–317 (2016).

21. D. Aujesky, M. J. Fine, The Pneumonia Severity Index: A Decade after the Initial Derivation and Validation. Clinical Infectious Diseases 47, S133–S139 (2008).

22. S. Papiris, A. Kotanidou, K. Malagari, C. Roussos, Clinical review: Severe asthma. Crit Care 6, 30–44 (2002).

23. K. E. J. Philip, E. Pack, V. Cambiano, H. Rollmann, S. Weil, J. O’Beirne, The accuracy of respiratory rate assessment by doctors in a London teaching hospital: a cross-sectional study. J Clin Monit Comput 29, 455–460 (2015).

24. G. H. P. Latten, M. Spek, J. W. M. Muris, J. W. L. Cals, P. M. Stassen, Accuracy and interobserver-agreement of respiratory rate measurements by healthcare professionals, and its effect on the outcomes of clinical prediction/diagnostic rules. PLoS One 14, e0223155 (2019).

25. Pneumonia (available at https://www.who.int/news-room/fact-sheets/detail/pneumonia).

26. ARIDA (Acute Respiratory Infection Diagnostic Aid) (available at https://www.unicef.org/innovation/arida).

27. A. M. Khan, A. O’Donald, T. Shi, S. Ahmed, E. D. McCollum, C. King, A. H. Baqui, S. Cunningham, H. Campbell, Accuracy of non-physician health workers in respiratory rate measurement to identify paediatric pneumonia in low- and middle-income countries: A systematic review and meta-analysis. J Glob Health 12, 04037.

28. M. J. Tobin, Why Physiology Is Critical to the Practice of Medicine: A 40-year Personal Perspective. Clinics in Chest Medicine 40, 243–257 (2019).

29. L. W. Andersen, K. M. Berg, M. Chase, M. N. Cocchi, J. Massaro, M. W. Donnino, American Heart Association’s Get With The Guidelines(®)-Resuscitation Investigators, Acute respiratory compromise on inpatient wards in the United States: Incidence, outcomes, and factors associated with in-hospital mortality. Resuscitation 105, 123–129 (2016).

30. A. D. Bedoya, N. A. Bhavsar, B. Adagarla, C. B. Page, B. A. Goldstein, N. R. MacIntyre, Unanticipated Respiratory Compromise and Unplanned Intubations on General Medical and Surgical Floors. Respir Care 65, 1233–1240 (2020).

