## Supplement - Materials and Methods for "Kinematic signature of high-risk labored breathing revealed by novel signal analysis"

**Supplementary File Contents:**

Section 1. Prototype Specifications (Page 2)

Section 2. Detecting non-respiratory motion artefact (Page 4)

Section 3. Validating respiratory rates in the exercise physiology laboratory (Page 11)

Section 4. Details of the 33 novel respiratory kinematic metrics (Page 13)

Section 5. Optimal metric selection and latent profile analysis (Page 18)

**Section 1. Prototype Specifications.**

We used MetaMotionR sensors by MbientLab (https://mbientlab.com/) to record thoraco-abdominal kinematics. These sensors contain a Bosch BMI160 inertial measurement unit (IMU) which records three orthogonal axes each (x, y, and z) of linear and angular acceleration signals in a synchronized manner. We programmed the IMUs to record each of the 6 signals at 100Hz. At each sensor location, therefore, we analyzed 600 inertial measurements per second (72,000 measurements per sensor per 2-minute recording).

We analyzed respiratory kinematics at 6 sensor locations. These included 3 upper rib-cage locations (midline sternal head and bilateral second rib in the midclavicular line), 2 lower rib-cage locations (bilateral eighth rib in the anterior axillary line), and one location on the abdomen (midline abdomen near umbilicus). We used MbientLab’s skin adhesive kit to secure sensors in place. The sampling rates were programmed identically across all sensors. Overall, we analyzed 432,000 inertial measurements per 2-minute recording (100 measurements per second x 120 seconds per recording x 6 signals per location x 6 locations).

Each of the 6 sensors established a Bluetooth Low Energy (BLE) link with a custom Android application running on a Samsung Galaxy A10e smartphone. The application implemented key functions such as (a) configuring sensors to a pre-specified setting (e.g. sampling rate of 100Hz), (b) communicating commands to start and stop signal recording, and (c) downloading recorded data for analysis.

Each sensor unit started and stopped a 2-minute recording interval according to its own microcontroller clock. However, we observed that the clock on one sensor unit drifted relative to that on another sensor unit during trials that lasted multiple hours. To combat this drift, we used the fact that the commands from the smartphone application to start the first and last recordings were transmitted simultaneously to each sensor. Since the timestamp of each command was measured to be accurate to 100ms, the maximum expected synchronization error between any two sensors of our prototype is 200ms. We used this information to scale the recorded time ranges such that time between phone commands would be constant across all six sensor units.

The clocks on the sensor chip controlling the 100Hz sampling were similarly imprecise; each sensor recorded a slightly different sample rate in each two minute interval even after correcting for the drift of the microcontroller clock. For example, if a given sensor chip’s clock was faster than the true 100Hz sampling, it could record 12001 samples in the two-minute period perceived by the microcontroller clock instead of the expected 12000 samples per recording. To manage this problem, we used the fact that each sensor always recorded the same number of samples per recording. For example, if a given sensor chip recorded 12001 samples in the first recording, then it recorded 12001 samples in every subsequent recording. This suggested that the difference in sampling rates was predictable and stable, and that the samples were evenly spaced. We therefore interpolated by resampling each stream by a rational factor (upsample by L, filter to 1/L, downsample by M to produce L/M interpolation). Interpolation aligned the samples across sensor units, but each sensor unit still had a different number of samples due to the differences in microcontroller clocks. The interval was cropped to the largest span that contained data from all sensor units. The result was a two-minute collection consisting of approximately 12,000 points each of three-axis linear and angular acceleration measurements, simultaneously sampled at exactly 100Hz across all sensor units.

**Section 2. Detecting non-respiratory motion artefact.**

We previously described a method to extract continuous respiratory rate signals from any stream of respiratory kinematic data *(1)*. However, any individual kinematic signal was prone to noise and the accuracy of the respiratory rate estimate was limited (95% limit of agreement of ± 7 breaths per minute) *(1)*. To address this problem, we combined the respiratory rate information in all 36 streams of kinematic data that we analyzed in this study (6 kinematic streams at each location x 6 locations). This work was based on the fact that any noise-free respiratory rate estimate would, by definition, be close to the true respiratory rate. Conversely, any noisy respiratory rate estimate would, by definition, not converge onto the true respiratory rate. This fact has two important implications. First, during a noise-free segment of the kinematic signals, a large proportion of the 36 respiratory rate estimates would converge on a particular RR. And second, when a large cluster of respiratory rate converged on a particular rate, then the centroid of the converging cluster would be a good estimate of the true respiratory rate. We used these considerations to create a data driven algorithm to detect noise and to improve the accuracy of respiratory rate estimation.

At any given point in time, we had 36 respiratory rate estimates. Comparing each rate to the other 35 rates at the same time, we obtained 630 pairwise differences in respiratory rate. We analyzed the distribution of these pairwise respiratory rate differences to arrive at a data-driven definition of “respiratory rate agreement.” We modelled the distribution of respiratory rate differences as being a composite of two separate distributions. The pairwise differences would have a certain expectation of typical performance if both respiratory rates in the pair measured the true respiratory rate, whereas it would have a different expectation if at least one respiratory rate in the pair was a noisy estimate.

Let $p^{ij}\left( x \right)$ denote the probability density function for the absolute pairwise difference $x=\left| {RR}_{i}-{RR}_{j} \right|$ between the respiratory rate (RR) estimated by sensors $i$ and $j$. Let $p_{a}^{ij}(x)$ be the probability of agreeing rates, and let $p_{d}^{ij}(x)$ be the probability of disagreeing rates. Interpolating between the two to gives the full distribution: $p^{ij}\left( x \right)=\alpha p_{a}^{ij}\left( x \right)+\left( 1-\alpha\right)p_{d}^{ij}\left( x \right)$ where $\alpha\in[0,1]$ is the linear interpolating factor. This amounts to a weighted average of the two distributions with weights of $\alpha$ and $(1-\alpha)$. Intuitively, the disagreeing distribution has a considerably fatter tail, as virtually all large differences are caused by large errors. We defined the closeness threshold of any two sensors to be the point of equal probabilities of error between the two parts. If an absolute difference is below the threshold, it is more probable that it is drawn from the distribution of both-valid rates; if above the threshold, it is more likely to that at least one sensor is noisy.

To model the overall distribution, samples of simultaneous pairwise absolute differences between sensors $i$ and $j$ were collected and visualized. Each histogram of related pairwise absolute differences steeply rises near $x=0$. We therefore modeled the agreeing rates as an exponential distribution:

$$p_{a}^{ij}\left( x \right)=\left\{ \begin{aligned} \lambda_{ij}e^{-\lambda_{ij}x}, &x\geq0 \\ 0, &x<0 \end{aligned} \right.$$

where $\lambda_{ij}$ is the exponential constant, equal to the reciprocal of the expected value of the distribution.

To model the disagreeing distribution, the distribution of unrelated pairwise differences was visualized for each pair. Unrelated pairwise differences were calculated by sampling rates from all collections and trials, shuffling the samples, and computing the difference of pairs of shuffled rates. This is not an exact model for the distribution of disagreeing rates, as some rates used are valid and not noise. Still, it provides a reasonable proxy, and as shown below, works perfectly well as a model for the remaining distribution.

The resulting histograms appeared normal; to model the data, the maximum likelihood estimate of normal distribution parameters were estimated. The distributions were fit as full normal distributions with means above and below zero; to compare with a single-sided exponential model, the absolute value of the mean was used. The new model with positive mean was folded at $x=0$ to capture the wrapped behavior of the absolute difference. Given positive mean $\mu_{ij}$ and variance $\sigma_{ij}^{2}$, the distribution of disagreeing rates can be approximated as:

$$p_{d}^{ij}\left( x \right)=\left\{ \begin{aligned} f_{n}\left( x;\mu_{ij},\sigma_{ij} \right)+ f_{n}\left( -x;\mu_{ij},\sigma_{ij} \right), &x\geq0 \\ 0, &x<0 \end{aligned} \right.$$

where $f_{n}$ is the normal probability density function.

With shapes for both functions and parameters for the disagreeing portion, the exact structure final probability distribution can be described:

$$p^{ij}\left( x \right)=\alpha p_{a}^{ij}\left( x \right)+\left( 1-\alpha\right)p_{d}^{ij}\left( x \right)$$

$$=\alpha\lambda_{ij}e^{-\lambda_{ij}x}+\left( 1-\alpha\right)\left[ f_{n}\left( x;\mu_{ij},\sigma_{ij} \right)+ f_{n}\left( -x;\mu_{ij},\sigma_{ij} \right) \right]$$

for non-negative $x$. The remaining task was to derive $\alpha_{ij}$ and $\lambda_{ij}$, which was again accomplished using MLE. This resulted in a complete distribution that fit the data very well, and notably better than a single exponential distribution (see Figure S1).

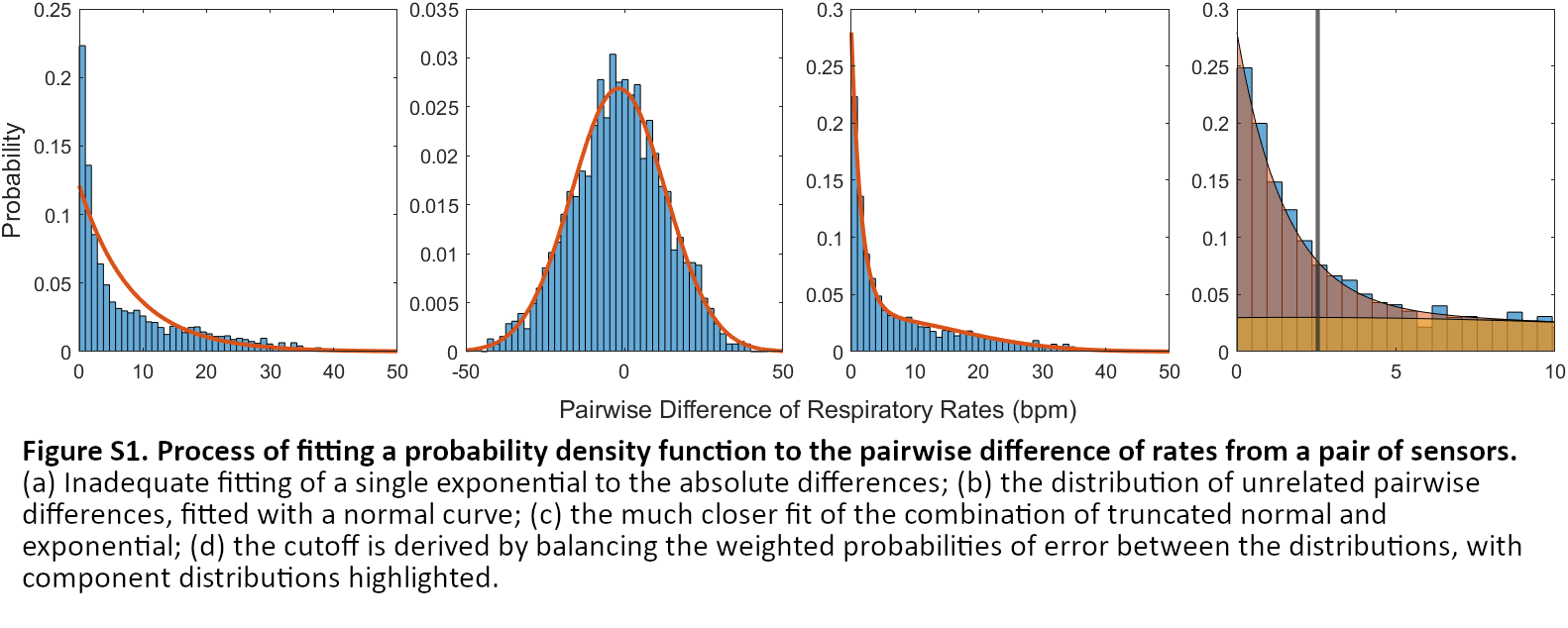

The final portion of the closeness analysis was the derivation of cutoffs, enabled by the modeling of contributing distributions. Error probability for the cutoff $c_{ij}$ is the cumulative distribution over the complementary domain:

$$p\left( error | agree \right)=p\left( error | disagree \right)$$

$$\alpha_{ij}\int_{c_{ij}}^{\infty} \lambda_{ij}e^{-\lambda_{ij}x}dx=\left( 1-\alpha_{ij} \right)\int_{c_{ij}}^{\infty} {[f}_{n}\left( x;\mu_{ij},\sigma_{ij} \right)+ f_{n}\left( -x;\mu_{ij},\sigma_{ij} \right)]dx$$

$$\alpha_{ij}\left( 1-F_{a}\left( c_{i,j} \right) \right)=\left( 1-\alpha_{ij} \right)F_{d}\left( c_{i,j} \right)$$

where $F_{i}(x)$ is the cumulative distribution function for $p_{i}\left( x \right)$.

Cutoffs were derived using the noisy data from the clinical trials, providing a realistic estimation of the noisiness of the distributions. The above process was repeated for each of the 630 unique pairs of different sensor streams.

The derived cutoffs can be used to label each pair of sensors at a given time to be classified as agreeing or disagreeing. It remains to be demonstrated how to find a single cluster of agreement from the pairs of rates. Consider an unweighted graph structure of the stream data, where each vertex $v_{i}$ represents a stream rate estimate. Vertices $v_{i}$ and $v_{j}$ are connected with an edge $e_{ij}$ if sensor streams $i$ and $j$ agree according to cutoffs (see Figure S2).

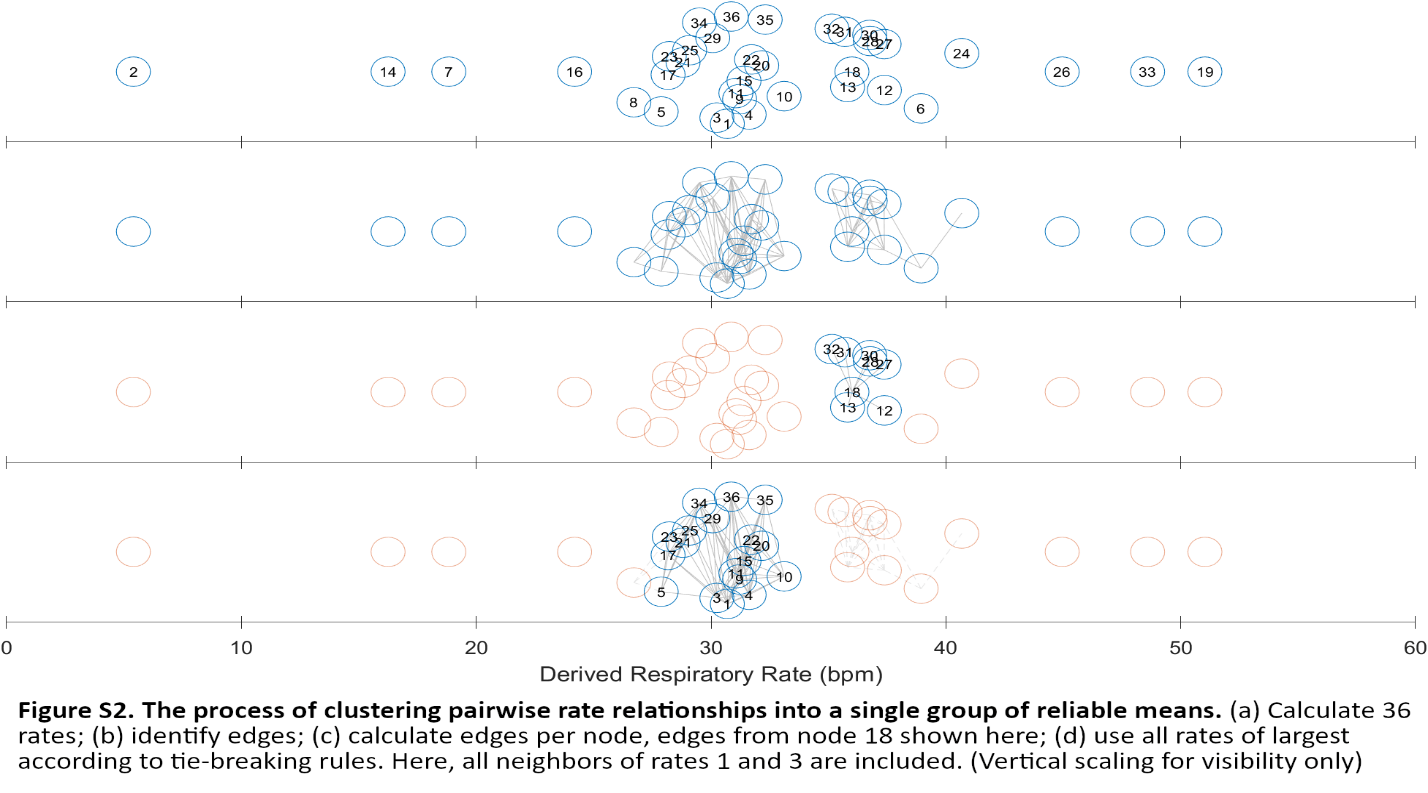

The traditional formulation of the largest cluster in a graph is known as a maximal clique, which is computationally difficult to extract. A more computable alternative is to group all vertices adjacent to the node with the most edges. This simpler statistic correlated with the max clique size in our data while minimizing the computation needed to find tying sets. Tying edge counts are often the product of a single cluster, with largely overlapping groups of constituent nodes. On the other hand, some signal groups produce two substantially different clusters of equal size. To break ties, the previous set of used points is maintained; all nodes with the maximum edge count that appeared in the previous set of nodes are considered equally good. The tie-breaking set of used nodes includes all rates adjacent to all reappearing max nodes. In the event that no max nodes appeared in the previous solution set, all max nodes and their adjacencies are considered to be useful. The previous node set is initialized as the empty set, so all max nodes are included on the initial time step.

With the agreeing cluster computed for each timepoint, the true rate can be calculated as the average of agreeing rates. The technique reliably finds the respiratory rate curve in a variety of stable and unstable breathing conditions.

Upon review, we observed that the quality of the agreement generally followed the number of signals used. We used this observation to develop a second layer of filtering on the respiratory rate time series, designed to detect useful data from the cluster information. We experimented with relative and absolute cutoffs for the count of rates used in each record. For the relative cutoffs, we tested preserving half the data in every record with a minimum level of agreement (12 rates, the lower quartile for average rates used in two minute segments). We observed a large disparity in quality between records where almost all the rates agreed and those where only a few agreed. Absolute cutoffs allowing all data above a certain threshold, on the other hand, produced fewer points per record, but the data selected was of much higher quality. We set a minimum absolute cutoff of at least 18 rates, reasoning that any cluster where half the rates agree is very likely to include the true rate. After reviewing higher thresholds, we selected exactly 18 rates, as increasing the cutoff eliminated substantially more good data than bad. Thus, the final respiratory rate series consisted of the cluster means, unevenly sampled such that all points used a cluster of at least 18 rates. Figure S3 demonstrates the removal of a noise artifact from the otherwise stable mean series.

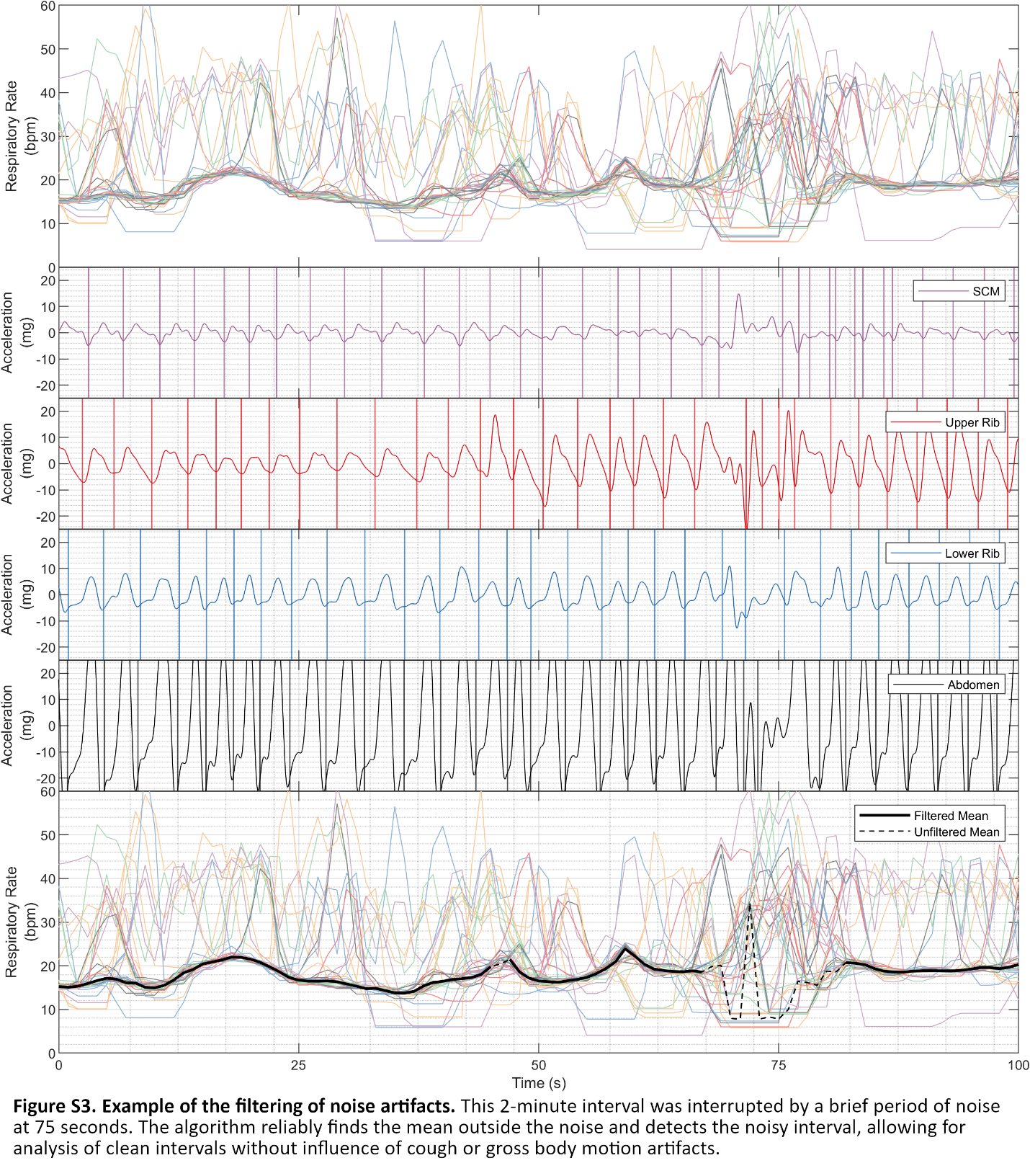

**Section 3. Validating respiratory rates in the Exercise Physiology Laboratory.**

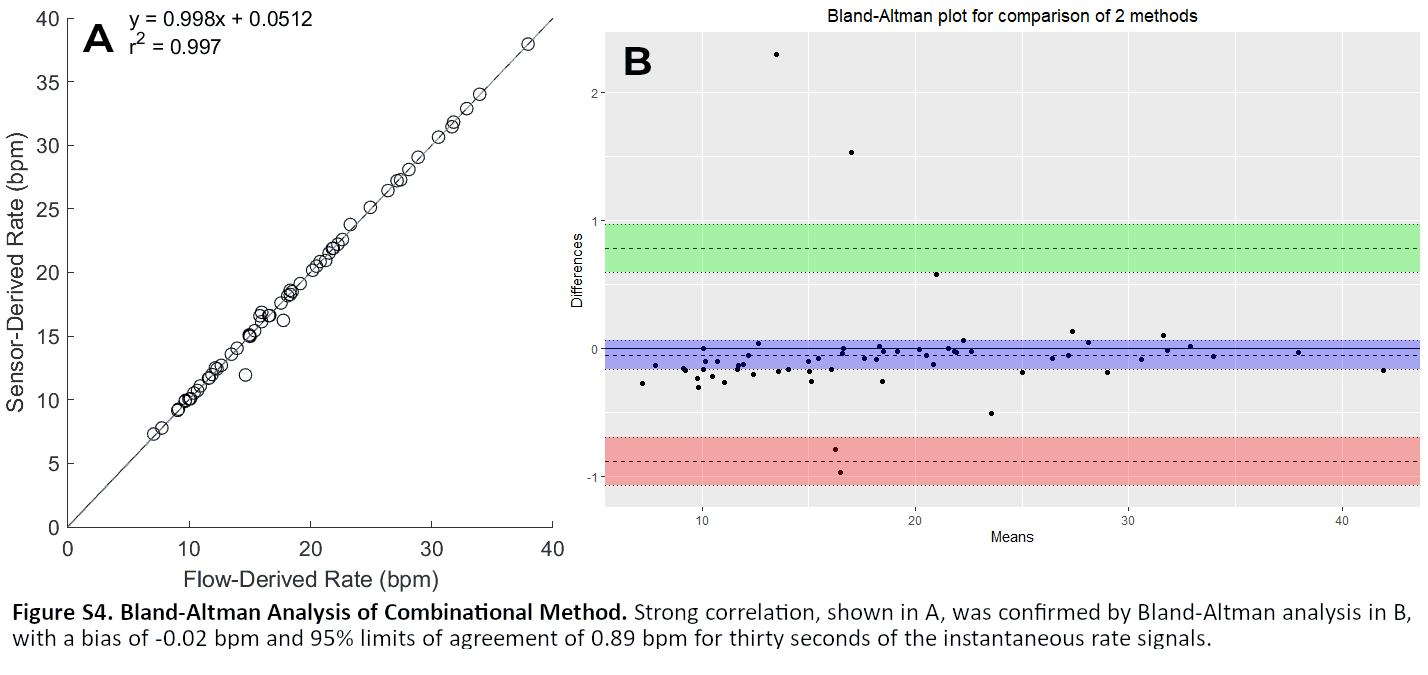
To determine the fidelity of the respiratory rate time series produced by the clustering process, we compared the clustered mean output to the respiratory rate series produced by the gold-standard flow streams from the exercise physiology dataset. Accuracy was evaluated across three segments from 20 trials, providing 60 total points of comparison between our algorithm and the ground truth from the flow data. Due to motion artifacts specifically from starting and stopping the breathing maneuvers, data for the accuracy calculations was cropped to be either 30 seconds or one minute long, starting 15 seconds into the recording.

Results can be seen in Figure S4. For the 30-second fidelity analysis, the coefficient of determination of the regression line between flow- and sensor-derived rate means was 0.997 (see Figure S4a), indicating a very close fit between the two mean rate values. The Bland-Altman analysis confirmed the stability of residuals, with a bias of -0.02 bpm and 95% limits of agreement at 0.89 bpm (see Figure S4b). Signals performed similarly over 60 seconds, with regression giving a coefficient of determination of 0.988 and Bland-Altman analysis producing a bias of 0.10 bpm and 95% limits of agreement of 1.55 bpm. Correlation analysis confirmed these results, with an average correlation of 0.89 for 30 seconds and 0.88 for 60 seconds.

Signals were evaluated by the root mean square (RMS) error and percent error of the instantaneous rate compared to the gold-standard flow rate signal, also derived via the Hilbert-Berger algorithm. The method achieves 3.7% error on average, with a minimum error of 0.76% on 30 seconds of data (4.5%, 1.2% for 60 seconds). The average RMS error was 0.89 bpm, with a best of 0.21 bpm (1.3 bpm, 0.30 bpm for 60 seconds). These results indicate that the method produces a mean time-series that is consistent with reference signals in spite of local variations in individual sensors and length of recording. Combined with the regression and Bland-Altman analysis, this evaluation demonstrates the utility of our methods for studying rate dynamics. Additionally, the information contained in the clustered streams provides evidence of the prevalence of motion noise artifacts, allowing analysis and removal of noise from body movement and coughing. The remaining error can be attributed to sensor noise, the minor lack of synchronization between the air flow and the kinematics of the chest, and uncertainty due to the re-synchronization of the flow data to the sensor data, as explained in *(13)*.

**Section 4. Details of the 33 novel respiratory kinematic metrics.**

We applied the newly derived cluster mean time series and associated filter information to calculate clinically sensible metrics across several categories: respiratory rate, respiratory rate variability; upper rib amplitude, lower rib amplitude, abdominal amplitude, ratio of upper rib to lower rib/abdominal amplitude, ratio of lower rib to abdominal amplitude, and variability of amplitude ratios.

For all metric calculations, the set of usable points was determined by the rate-agreement filter. For respiratory rate and rate variability, this filtered series was all points in the clustered rate series with at least 18 rates. We set a minimum acceptability criterion of 30 noise-free points. 582 of the 790 (74%) of the recordings met this criterion. Metrics were calculated for the 582 acceptable recordings. The amplitude ratio were calculated using the 100Hz acceleration signals, so the usable acceleration series consisted of the union of one-second intervals centered on the filtered points from the respiratory rate time series.

For each category, multiple metrics were calculated to determine the optimal method for extracting physiological information. Typical respiratory rate was calculated as the average of usable points. Respiratory rate variability was calculated first as the standard deviation and coefficient of variation of the filtered respiratory rate time series, but we observed that the dependence on outliers was strong. To improve the integrity and robustness of the variability metric, we also considered sequential differences of contiguous segments of data spaced one, two, or three samples apart. For each spacing, we calculated the mean, median, and maximum of the set of differences for a total of eleven variability metrics.

To extract acceleration-based metrics, we calculated acceleration magnitude time series by combining the individual dimensions of the 3-axis acceleration signal. We processed each acceleration stream for an envelope. While the amplitude function from the analytic representation provides an envelope, the wide frequency band of breathing signals introduces artifacts that are incorrectly expressed as oscillations in the amplitude at the same rate as breathing. The resulting envelope is too noisy for magnitude analysis without further smoothing. Still, the extrema of a signal are well-detected by the analytic representation. To ensure a robust smoothing procedure, the maxima and minima of each signal were combined into individual upper and lower envelopes using an uneven upsampling process. The original time vector was used as the target time vector; each time not containing a landmark was filled with the amplitude of the nearest landmark. This step wave was filtered such that the cutoff frequency was equal to the reciprocal of the longest time between landmarks. This guaranteed that frequencies appearing in the envelope were accurately reconstructed while staying robust to potentially long gaps for noisy records. The method improves upon the stability of the analytic envelope while avoiding the volatility of spline interpolation, which produces unusable results for signals with sparse landmarks due to low amplitude and even inverted envelopes where one swings past the other. The instantaneous amplitude was calculated as the difference between the upper and lower envelopes. The final acceleration envelopes ${{}^{e}a}_{sd}$ for each sensor $s$ and dimension $d$ were combined with a Euclidean norm into the sensor acceleration magnitude, ${{}^{e}a}_{s}$:

$${{}^{e}a}_{s}\left( t \right)=\sqrt{{{}^{e}a}_{sx}^{2}\left( t \right)+{{}^{e}a}_{sy}^{2}\left( t \right)+{{}^{e}a}_{sz}^{2}\left( t \right)}$$

Motion signals for the four vertical positions (SCM, upper rib, lower rib, and abdomen) were derived by combining bilateral signals with a simple average. While lateral differences in breathing motion are documented (i.e. flail chest), they are relatively unusual and almost always obvious to the naked eye. Thus, bilateral measurement was used to reinforce the reliability of measurements:

$${{}^{e}a}_{UR}\left( t \right)=\frac{{{}^{e}a}_{L2}\left( t \right)+{{}^{e}a}_{R2}\left( t \right)}{2}$$

$${{}^{e}a}_{LR}\left( t \right)=\frac{{{}^{e}a}_{L8}\left( t \right)+{{}^{e}a}_{R8}\left( t \right)}{2}$$

This produced four regional signals, for the SCM, upper ribs, lower ribs, and abdomen. The average amplitude was calculated over all one-second regions containing a filtered mean sample, producing four metrics.

From the four regional signals, we calculated a ratio between the upper signals and lower signals, denoting this to be the index of Recruitment of Accessory Muscles (RAM). For the initial version, each pair of regions were combined by adding them together:

$$RAM\left( t \right)=\frac{{{}^{e}a}_{SCM}\left( t \right)+{{}^{e}a}_{UR}\left( t \right)}{{{}^{e}a}_{LR}\left( t \right)+{{}^{e}a}_{Abd}\left( t \right)}$$

We hypothesized that the relationship could be improved by selecting signals to intentionally exacerbate the metric. By definition, RAM increases for larger upper compartment magnitudes and smaller lower compartment magnitudes. Given that different subjects may vary their recruitment of each component signal, the resulting upper and lower magnitudes may unnecessarily dilute the effect of upper muscle recruitment. To combat this effect, opportunistic RAM (opRAM) was developed as a companion metric. In contrast to RAM, opRAM is calculated using the minimum of the lower ribs and abdomen as the lower signal, and similarly the maximum of the SCM and upper ribs as the upper signal:

$$opRAM\left( t \right)=\frac{\max\left( {{}^{e}a}_{SCM}\left( t \right),{{}^{e}a}_{UR}\left( t \right) \right)}{\min\left( {{}^{e}a}_{LR}\left( t \right),{{}^{e}a}_{Abd}\left( t \right) \right)}$$

A common transformation for extracting information from ratio quantities is to use the logarithm of the actual ratio to compare signals, as is done in with signal-to-noise ratios and decibel units. Whereas a linear relationship prescribes an absolute meaning to the ratio unit, the logarithm can better describe and compare the behavior of the signal when multiplicative factors have more meaning and makes resulting measures invariant to the magnitude of the underlying signals. Thus, the logarithmic transformations, $\log\left( RAM\left( t \right) \right)$ and $\log\left( opRAM\left( t \right) \right)$, have the potential to extract more meaning from the comparison of the rib cage compartments. From each of these four series, the mean was calculated, producing four metrics.

To measure fluctuations in lower compartment contribution, we calculated the RA ratio as the ratio between the upper ribs to the abdomen magnitude, similarly to RAM:

$$RA\left( t \right)=\frac{{{}^{e}a}_{LR}\left( t \right)}{{{}^{e}a}_{Abd}\left( t \right)}$$

The logarithm of the ratio was again used as an additional processing step to extract variations in ratio quantities. Summarizing RA activity was more difficult; as opposed to the consistency of RAM, RA is characterized by a transient effect. To this end, the mean, standard deviation, maximum, minimum, and range signals were initially considered for each two-minute collection, producing four metrics. Similarly to the variance of breathing rate, a lack of confidence in the ability of standard deviation to capture the variation of interest led to the development of sample-difference metrics. Thus, the log RA ratio signal was downsampled to 1 Hz to match the respiratory rate signal, and first differences of multiple spacings (1, 2, and 3 seconds) were taken, producing nine additional metrics.

Again, from each of these collections, the average, median, and the maximum were calculated and included with the other summary metrics for further analysis.

| Table S1. Summary of Metrics Produced for Analysis | | |
| --- | --- | --- |
| Metric Type | Number of Metrics | Metric Names |
| Respiratory Rate Variability | 11 | RR Standard Deviation  RR Coefficient of Variation  RR Maximum n-second Difference (n = 1,2,3)  RR Minimum n-second Difference (n = 1,2,3)  RR Mean n-second Difference (n = 1,2,3) |
| Regional Magnitudes: Upper chest motion | 2 | Sternocleidomastoid Mean Amplitude  Upper Rib Mean Amplitude |
| Regional Magnitudes: Lower chest motion | 1 | Lower Rib Mean Amplitude |
| Regional Magnitudes: Abdominal motion | 1 | Abdomen Mean Amplitude |
| Amplitude ratios: Upper rib to lower rib and abdomen  (Recruitment of Accessory Muscles) | 4 | RAM Mean  opRAM Mean  log RAM Mean  log opRAM Mean |
| Amplitude ratios: Lower rib to abdomen | 4 | Mean ratio  Mean Log ratio  Maximum Log ratio  Minimum Log ratio |
| Variability of amplitude ratio: lower rib to abdomen  (Respiratory Alternans) | 10 | RA Standard Deviation  RA Maximum n-second Difference (n = 1,2,3)  RA Minimum n-second Difference (n = 1,2,3)  RA Mean n-second Difference (n = 1,2,3) |

**Section 5. Optimal metric selection and latent profile analysis.**

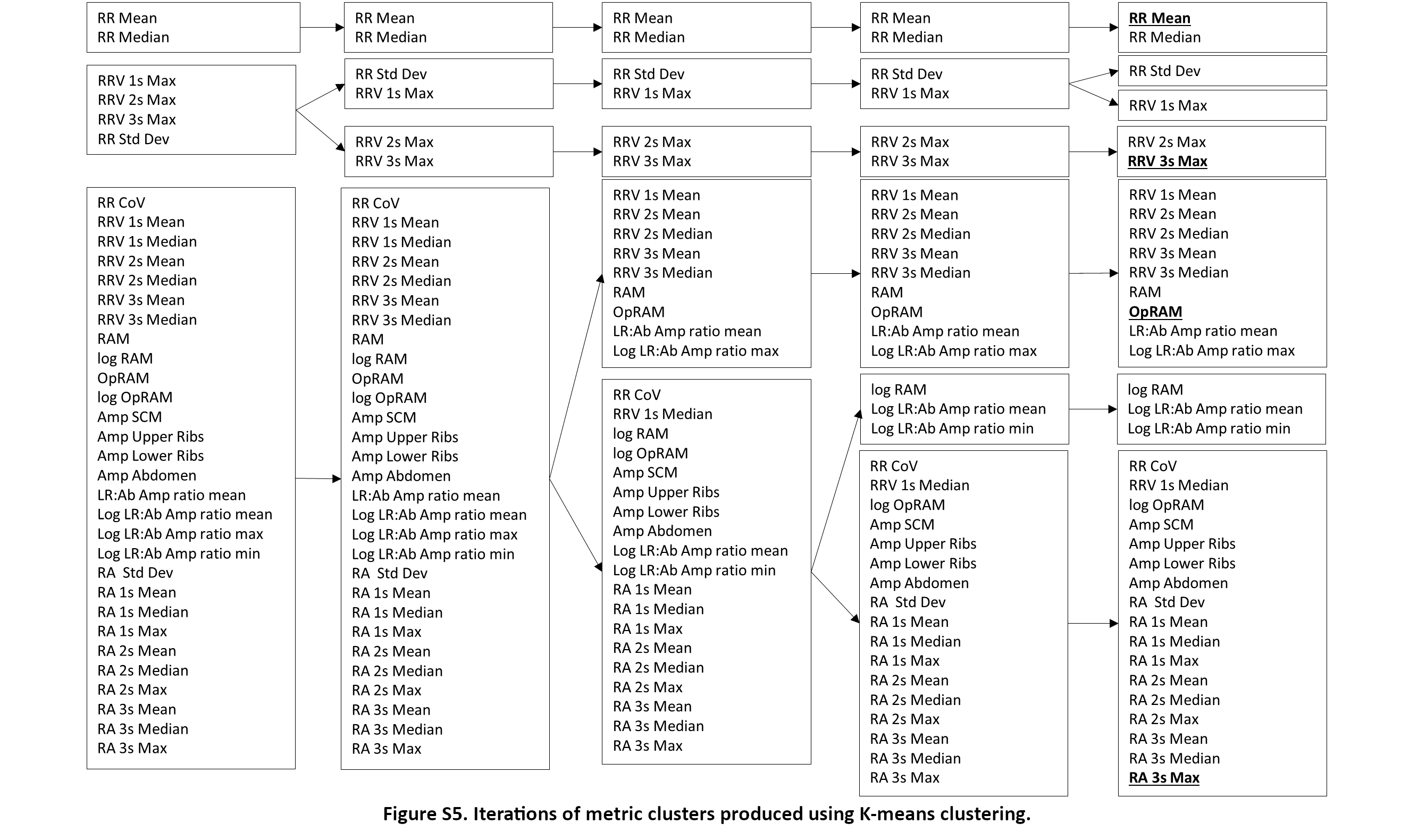
We selected the mean respiratory rate because of its indisputable importance as a conventional vital sign. To determine the minimum number of novel metrics required to optimally represent the respiratory kinematic variability in our dataset, we used unsupervised k-means clustering to identify clusters of metrics across the 582 recordings.

By looking for an “elbow” in the Within Sum of Squares (WSS) plot, we determined that selecting three novel metrics from separate clusters would be the optimal choice for our dataset, in addition to mean respiratory rate.

Using clinical domain expertise, we selected metrics that represented respiratory rate variability (short breath intervals alternating with long ones), recruitment of accessory muscles (magnitude of upper rib excursions relative to that of lower ribs and abdomen) and respiratory alternans (rib dominant breaths alternating with abdomen dominant ones). We analyzed the scoring patterns across the selected respiratory kinematic measures using latent profile analysis (LPA).

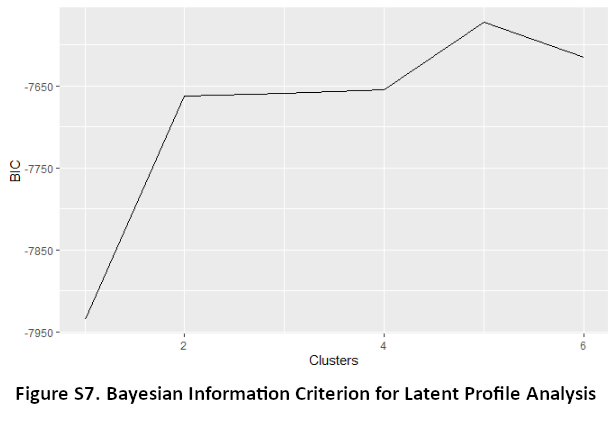

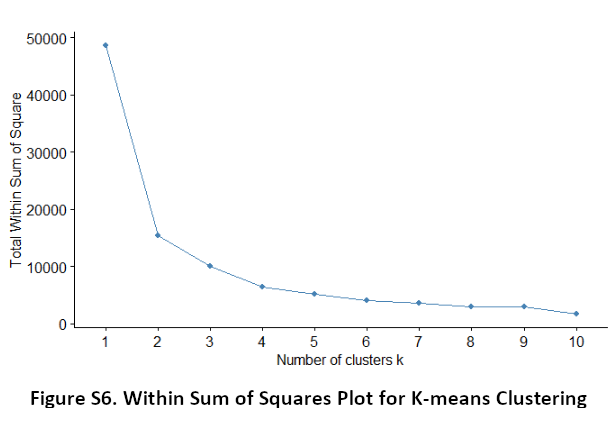
We aimed to minimize the Bayesian Information Criterion (BIC) to select optimal number of profiles with two constraints: (a) avoid small profiles that contained fewer than 58 recordings (10% of the 582 recordings), and (b) BIC improvement of at least 10 points. The lowest BIC was associated with 5 profiles, but 2 of the profiles had fewer than 58 recordings. And, there was no significant BIC difference between 2, 3, and 4 profiles. As such, we decided that our data contained two latent profiles.
